# A pharmacometric grey zone reconciles high metronidazole resistance rates with bismuth quadruple therapy efficacy in *Helicobacter pylori*

**DOI:** 10.64898/2026.06.21.26356170

**Authors:** Chunmeng He, Huan Wang, Xinbo Xu

## Abstract

**Background:** Metronidazole (MET) resistance in *Helicobacter pylori* (*H. pylori*) exceeds 50–60% globally, yet MET-containing bismuth quadruple therapy (BQT) achieves >90% eradication in MET-resistant infections. We hypothesise this discordance stems from a structural limitation of two-fold dilution: a pharmacometric grey zone between the 128 and 256 µg/mL breakpoints where treatable isolates are systematically misclassified as high-level resistance.

**Methods:** In a real-world cohort of 4610 treatment-naïve children (2019–2024), checkerboard assays determined the bismuth–MET synergy factor (SF). Population PK/PD modelling simulated gastric MET exposure (AUC_0-24_/MIC≥70) at 1 g, 1·5 g and 2 g daily doses, incorporated bismuth SF, and estimated MIC coverage against the European Committee on Antimicrobial Susceptibility Testing (EUCAST) population distribution. Findings were validated against published adult trials.

**Findings:** 56·4% (485/860) of isolates were MET-resistant (MIC>8 µg/mL). At ≤1 g/day MET, BQT offered no advantage over triple therapy for resistant infections. At 1·5 g/day, the synergy threshold was surpassed, achieving >90% eradication in MET-resistant infections, with efficacy independent of baseline susceptibility status. PK/PD modelling showed 1·5 g/day MET plus bismuth (median SF=8) extended coverage to ∼180 µg/mL. To align modelled coverage with the >90% eradication rate, the dominant subpopulation of nominally high-resistant isolates must have a geometric mean MIC well below 256 µg/mL (centred near 150 µg/mL), consistent with a mixture of two log-normal subpopulations and confirming systematic upward misclassification by two-fold dilution in the grey zone.

**Interpretation:** This grey zone is a structural blind spot of two-fold dilution, not random measurement error. The ±1-dilution reproducibility limit renders the interval inherently unresolvable. We propose a three-tier strategy: omit routine MET susceptibility testing for treatment-naïve patients on optimised BQT; reserve categorical testing for BQT failures; and explore genomic/metabolomic biomarkers beyond MIC paradigms in this enriched population.

**Funding:** None.

**Research in context:** *Evidence before this study:* Global metronidazole (MET) resistance in *Helicobacter pylori* (*H. pylori*) now exceeds 50–60%, posing a major challenge to first-line eradication therapy. Paradoxically, MET-containing bismuth quadruple therapy (BQT) consistently achieves >90% eradication in MET-resistant infections — a discordance that remains mechanistically unexplained. Previous work has established that MET resistance arises from stepwise loss of nitroreductase function, producing the trimodal MIC distribution formalised by the European Committee on Antimicrobial Susceptibility Testing (EUCAST). Bismuth has been shown to synergise with MET *in vitro*, but the concentration dependence of this interaction and its clinically relevant dose threshold remain poorly defined, leaving MET dosing in BQT largely empirical. While the ±1 two-fold dilution reproducibility limit of standard antimicrobial susceptibility testing (AST) is well documented, the structural blind spot at the 128–256 µg/mL interval has never been systematically characterised, nor has this methodological limitation been linked to the clinical resistance–efficacy paradox. No prior study has integrated real-world clinical outcomes, *in vitro* synergy data, and pharmacokinetic/pharmacodynamic (PK/PD) modelling to reconcile the gap between reported resistance rates and observed treatment success.

*Added value of this study:* This study identifies and quantitatively characterises a pharmacometric grey zone between the 128 and 256 µg/mL breakpoints, demonstrating that standard two-fold dilution systematically misclassifies treatable isolates in this interval as high-level resistance—providing a parsimonious explanation for the longstanding clinical–microbiological discordance. We further show that the nominal high-resistance peak is not a unimodal log-normal distribution, but a mixture of two distinct subpopulations: a dominant grey-zone subpopulation comprising ∼96% of nominally high-resistant isolates, with a true geometric mean MIC near 150 µg/mL, and a small minority of genuine high-level resistance. This reframes the understanding of “high-resistance” as a heterogeneous category rather than a single phenotypic state. Additionally, we define a clear dose threshold for bismuth–MET synergy: 1·5 g/day MET achieves the additive window and covers the vast majority of resistant isolates, providing a PK/PD rationale for empiric dosing and resolving prior empirical uncertainty. Finally, we propose a practical three-tier AST strategy that directly informs clinical decision-making and prioritises enriched populations for future biomarker discovery. While hypothesis-generating in design, the integration of real-world clinical data, *in vitro* synergy assays, PK/PD modelling, and external validation creates a logically consistent framework robust to key sensitivity analyses.

*Implications of all the available evidence:* Clinically, these findings challenge the utility of routine pre-treatment MET susceptibility testing for treatment-naïve patients. Optimised BQT with 1·5 g/day MET covers most nominally resistant isolates, meaning categorical AST results rarely alter first-line management, and clinicians should not abandon MET solely on the basis of a “high-resistance” classification. For paediatric populations, a 30 mg/kg/day dose is pharmacodynamically justified, balancing efficacy and safety. For diagnostics and research, the existence of a structural grey zone highlights fundamental limitations of current two-fold dilution AST frameworks, which reduce continuous functional resistance into discrete, potentially misleading categories. Future resistance studies should move beyond binary susceptible/resistant classifications to account for the graded, stepwise nature of nitroreductase impairment. Biomarker discovery efforts should focus on the enriched population of patients who fail optimised BQT despite nominal high resistance, rather than unselected cohorts, to maximise the likelihood of identifying clinically actionable signatures. Finally, gastroretentive MET formulations warrant further investigation as a means to further increase intragastric exposure, expand coverage of the grey zone, and reduce systemic toxicity.

## Introduction

*Helicobacter pylori* remains the primary aetiological agent of peptic ulcer disease and gastric cancer.^1^ As clarithromycin (CLA) resistance exceeds 15% globally, bismuth quadruple therapy (BQT) has become first-line treatment.^2–4^

Metronidazole (MET) is a prodrug activated by nitroreductase RdxA; resistance arises primarily from *rdxA* inactivation, with *frxA* mutations elevating resistance further.^5^ MET resistance now surpasses 60% in many regions,^2,6^ yet MET-containing BQT consistently achieves >90% eradication even in MET-resistant infections — a discordance not observed for CLA or levofloxacin (LEV).^2,3,7,8^ This points to a limitation of conventional two-fold dilution MIC testing, which cannot resolve the graded loss of nitroreductase function underlying the European Committee on Antimicrobial Susceptibility Testing (EUCAST) trimodal distribution.^9^

Standard antimicrobial susceptibility testing (AST) collapses all isolates with MIC≥256 µg/mL into a single bin, potentially misclassifying treatable isolates in the 128–256 µg/mL pharmacometric grey zone as high-level resistance. The ±1-dilution reproducibility limit renders the interval inherently unresolvable, and refined gradient testing lacks inter-laboratory reproducibility for *H. pylori* (ISO 20776-1). This discretisation artefact has been overlooked, and MET dosing in BQT remains empirical. In Chinese adult populations, this uncertainty has led to premature abandonment of MET,^10^ whereas paediatric guidelines still rely on it.^4,11^ Bismuth has intrinsic anti-*H. pylori* activity and may synergise with MET.^12^ We hypothesised that bismuth–MET synergy is concentration-dependent: exceeding an intragastric MET “additive window” renders most high-level resistance pharmacodynamically irrelevant, and the 128–256 µg/mL interval constitutes a structural grey zone where two-fold dilution systematically overestimates true resistance. We integrate real-world paediatric data, checkerboard assays, and population PK/PD modelling to define the synergy threshold and quantify the grey zone.

## Methods

### Study design and participants

This single-centre retrospective cohort enrolled treatment-naïve children (6–18 years) with first-diagnosed *H. pylori* infection at Fudan University Children’s Hospital (2019–2024). Exclusions included prior eradication therapy, recent antibiotic/PPI/bismuth use, immunodeficiency, or chronic gastrointestinal comorbidities. The study followed STROBE guidelines and was approved by the institutional ethics committee; guardians provided written informed consent.

### Procedures and outcomes

Patients received 14-day weight-based eradication regimens per the 2022 Chinese paediatric consensus,^11^ stratified into BQT, concomitant therapy (CT), or triple therapy (TT) (supplementary table S1). MET was administered at 20 mg/kg/day in two divided doses, with a maximum daily dose of 1000 mg (500 mg twice daily) per guideline specifications. Across the 6–18-year-old study cohort, actual total daily MET doses ranged from approximately 400 mg in younger children (≈20 kg body weight) to 1000 mg in adolescents weighing ≥50 kg. For translational framing, the adult reference daily doses of 1 g, 1·5 g and 2 g correspond to paediatric weight-based doses of ∼20 mg/kg/day, ∼30 mg/kg/day and ∼40 mg/kg/day respectively, when normalised to a 50 kg reference adolescent body weight.

The primary endpoint was eradication success, defined as negative ^13^C-urea breath test or rapid urease test ≥4 weeks post-therapy, following a 2-week PPI/antibiotic/bismuth washout.^3,11^

### Microbiological characterisation

Gastric biopsies were cultured and identified by standard methods (Supplementary Methods). AST used Etest (BIO-KONT) per EUCAST guidelines (v14.0, 2024).^13^ Resistance breakpoints: MET>8 µg/mL, CLA>0·25 µg/mL, amoxicillin (AMO)>0·125 µg/mL, LEV>1 µg/mL (surveillance only).

Etest’s continuous gradient can technically resolve intermediate values within the between the 128 and 256 µg/mL breakpoints range, but routine laboratory practice rounds these to the nearest two-fold dilution for EUCAST reporting. This operational convention, compounded by the ±1 two-fold dilution reproducibility limit of broth microdilution (ISO 20776-1), renders the between the 128 and 256 µg/mL breakpoints interval operationally indistinguishable—a structural blind spot, not merely a technical limitation.

### In vitro synergy

Checkerboard microdilution assays used *H. pylori* ATCC 43504 and four MET-resistant clinical isolates (MIC 32 to >256 µg/mL). Interactions were classified by fractional inhibitory concentration index (FICI; Supplementary Methods). Synergy factor (SF) was defined as MET MIC alone divided by MIC with 0·5×bismuth MIC.^12^ The additive window was defined as FICI≤1·0;^14^ its lower boundary informed PK/PD modelling. All assays were performed in duplicate across three independent experiments.

### Population PK/PD modelling

Plasma and gastric fluid MET concentrations were simulated using a validated population PK model.^15^ The pharmacodynamic target was AUC_0-24_/MIC≥70.^16^ Three adult reference daily doses (1 g, 1·5 g, 2 g) were modelled to define the concentration threshold for bismuth–MET synergy; gastric fluid AUC0-24 was derived by multiplying plasma AUC by 4·5 (gastric-juice-to-plasma ratio under PPI therapy).^15^ These adult dose levels were selected as the calibration standard to align with published clinical efficacy data and EUCAST population MIC distributions. Since the PK/PD target (AUC0-24/MIC ≥ 70) is exposure-based and age-independent, the 1·5 g/day adult threshold translates to a paediatric equivalent dose of approximately 30 mg/kg/day when adjusted for body weight, consistent with allometric scaling principles.

Bismuth synergy was incorporated as: effective MIC=observed MIC/SF. Primary analysis used median SF=8, with sensitivity analyses at SF=2, 4, 16. Maximum coverable MIC=(gastric AUC_0-24_/70)×SF.

MIC coverage was calculated against the EUCAST trimodal distribution (n=11,168 isolates: Peak 1 ≤8 µg/mL, Peak 2 16–128 µg/mL, Peak 3 ≥256 µg/mL).^9^ To quantify the grey zone, we reconciled modelled coverage with observed >90% eradication in MET-resistant infections treated with 1·5 g/day BQT. Stepwise derivation of the bimodal Peak 3 model is in Supplementary Methods. External validation used published adult trial and registry data.^7,8^

### Statistical analysis

Categorical variables were compared by χ^2^ or Fisher’s exact test; continuous variables by t-test or Mann–Whitney U test. Propensity score matching (PSM) (1:1, caliper 0·2) adjusted for age, sex, endoscopic findings, and treatment year (supplementary figure S1). Multivariable logistic regression identified independent predictors of eradication success. Sensitivity analyses assessed loss to follow-up impact. A two-sided p<0·05 was considered significant. Analyses used SPSS 20.0 (IBM).

## Results

### Study population and resistance profiles

Of 4,610 eligible children, 1,844 (40·0%) completed post-treatment assessment and formed the efficacy cohort (figure 1). Resistance profiles were comparable between followed and loss to follow-up patients (supplementary table S2), supporting representativeness. BQT was the predominant regimen (54·0%), followed by TT (43·6%) and CT (2·4%).

**Figure 1.**
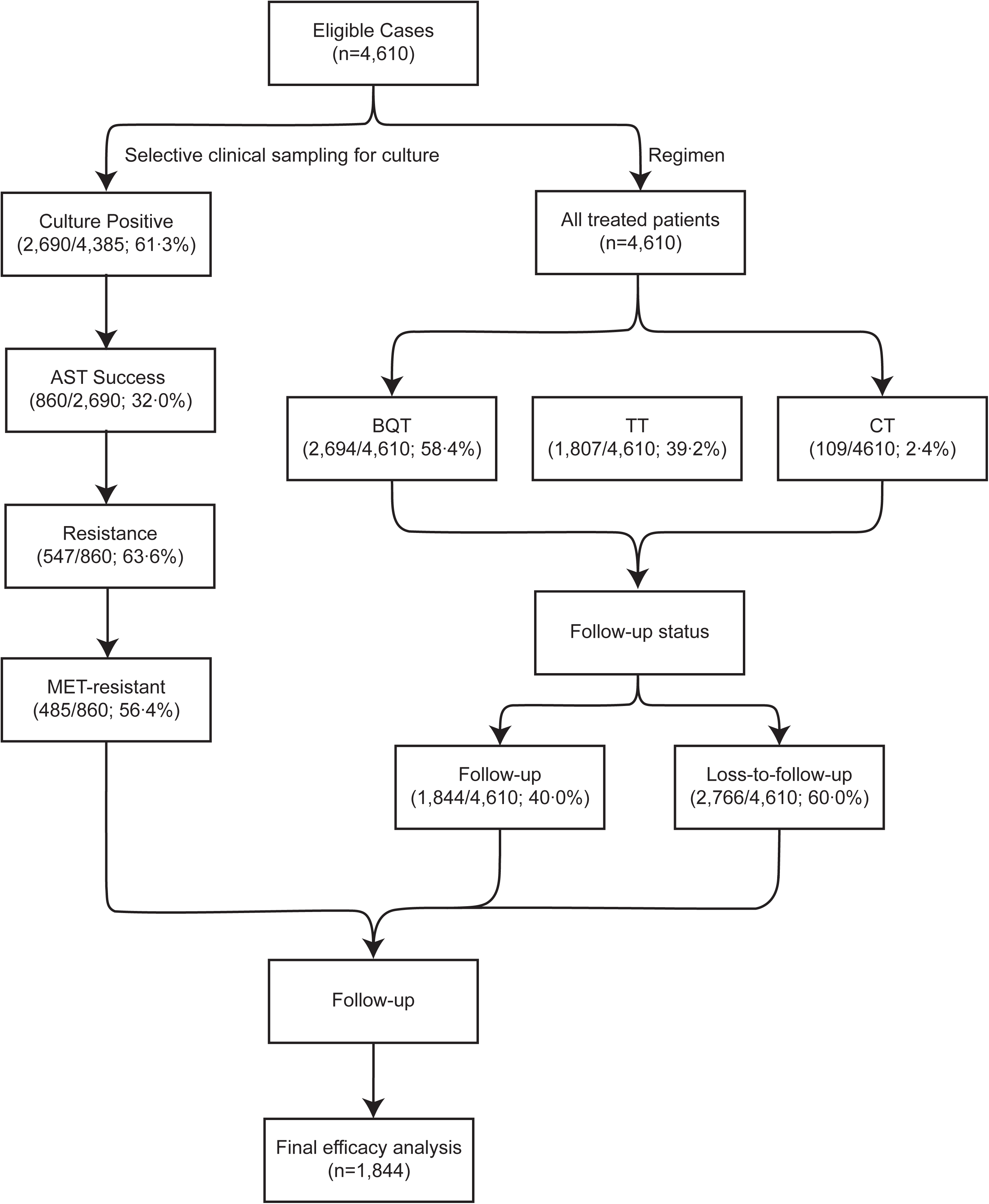
Flow chart of patient enrolment, microbiological testing, treatment allocation and follow-up. This single-centre retrospective cohort included 4610 treatment-naïve paediatric patients with confirmed *Helicobacter pylori* infection. Two parallel analytical pathways are shown: microbiological susceptibility testing (left) and clinical treatment outcome assessment (right). Of 4385 patients who underwent selective clinical sampling for bacterial culture, 2690 (61·3%) yielded positive cultures. Valid antimicrobial susceptibility testing (AST) was achieved in 860 isolates (32·0% of culture-positive samples); 547 isolates (63·6%) were resistant to at least one antibiotic, of which 485 (56·4%) were metronidazole-resistant. All eligible patients received first-line eradication therapy: 58·4% received bismuth quadruple therapy (BQT), 39·2% received triple therapy (TT), and 2·4% received concomitant therapy (CT). At post-treatment efficacy assessment, 2766 patients (60·0%) were lost to follow-up; 1844 patients (40·0%) with complete outcome data constituted the final per-protocol efficacy analysis cohort. Abbreviations: AST, antimicrobial susceptibility testing; BQT, bismuth quadruple therapy; CT, concomitant therapy; MET, metronidazole; TT, triple therapy.

Among 860 isolates with successful AST, overall resistance to any antibiotic was 63·6% (95% CI 60·4–66·8%): MET 56·4% (53·1–59·7%), CLA 26·7% (23·8–29·9%), AMO 2·7% (1·8–4·0%), LEV 0·5% (0·2–1·2%) (figure 2A).

**Figure 2.**
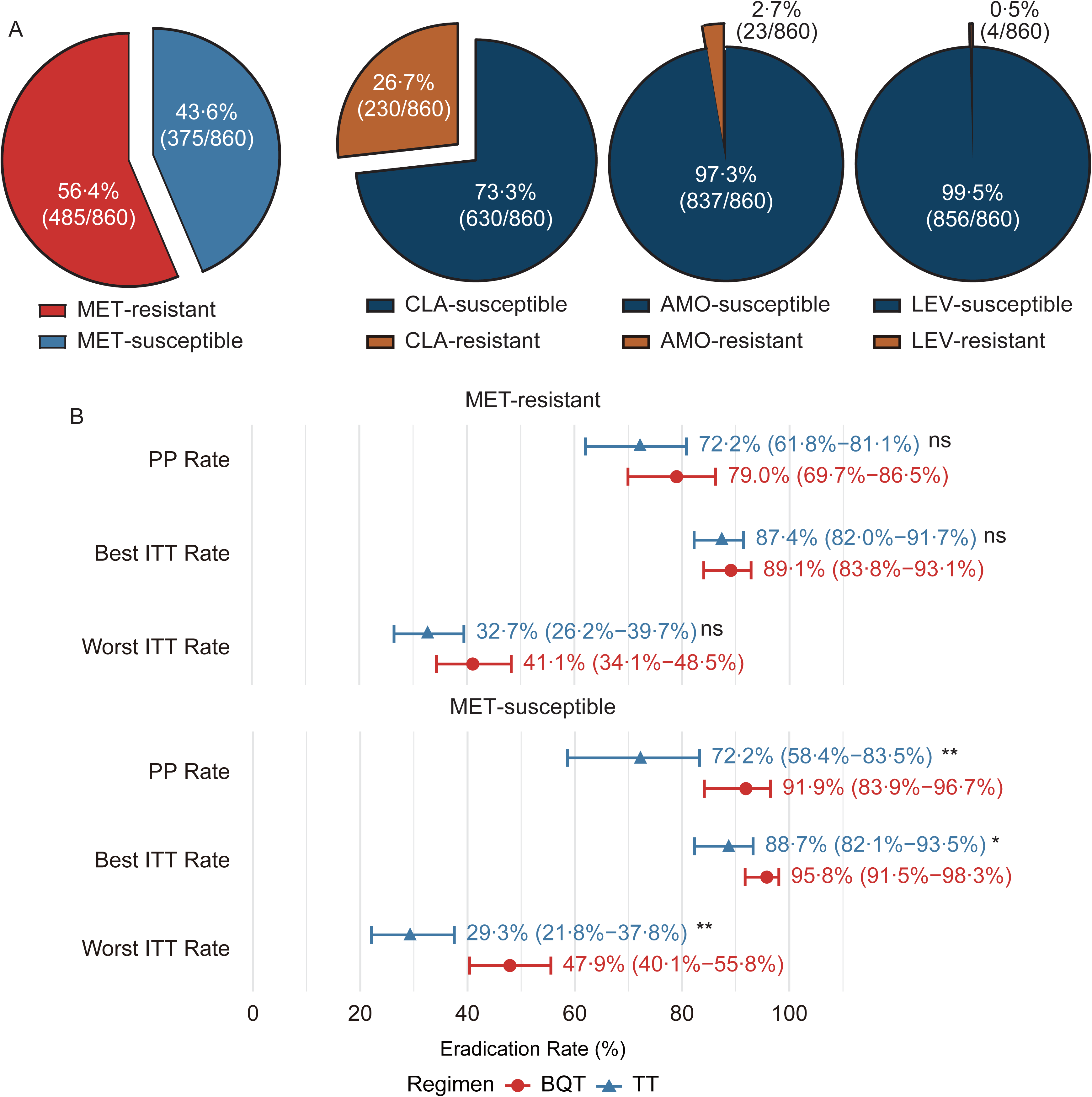
Antibiotic resistance profiles and eradication rates of different treatment regimens. (A) Pie charts illustrating the resistance and susceptibility rates to metronidazole (MET), clarithromycin (CLA), amoxicillin (AMO), and levofloxacin (LEV) among 860 isolates. Red and blue segments represent MET-resistant and MET-susceptible strains, respectively. For CLA, AMO, and LEV, dark blue and brown segments denote susceptible and resistant strains, respectively. Percentages and corresponding case numbers (e.g., 56·4%, 485/860) are annotated on the charts. (B) Forest plots comparing the eradication rates of bismuth quadruple therapy (BQT, red circles) and triple therapy (TT, blue triangles) in MET-resistant and MET-susceptible populations. The outcomes include per-protocol (PP) rates, best-case intention-to-treat (ITT) rates, and worst-case ITT rates. Error bars indicate 95% confidence intervals (CIs). “ns” denotes no significant difference, while * and ** indicate statistical significance at p<0·05 and p<0·01, respectively. The x-axis represents the eradication rate (%). Abbreviations: MET, metronidazole; CLA, clarithromycin; AMO, amoxicillin; LEV, levofloxacin; PP, per-protocol; ITT, intention-to-treat; BQT, bismuth quadruple therapy; TT, triple therapy; CI, confidence interval.

### Dose-dependent synergy threshold

After PSM, across per-protocol (PP), best-case intention-to-treat (ITT) and worst-case ITT sensitivity analyses, BQT showed no significant eradication benefit over TT among MET-resistant cases at ≤1 g/day MET (all p≥0·05) (figure 2B, table 1, supplementary table S3). At ≥1·5 g/day, BQT achieved >90% eradication regardless of susceptibility.^2,3,7,8^ In multivariable analysis, MET resistance was not an independent predictor of failure (adjusted OR 0·6, 95% CI 0·33–1·07, p=0·087), whereas BQT predicted success (adjusted p=0·016) (table 2).

**Table 1.**
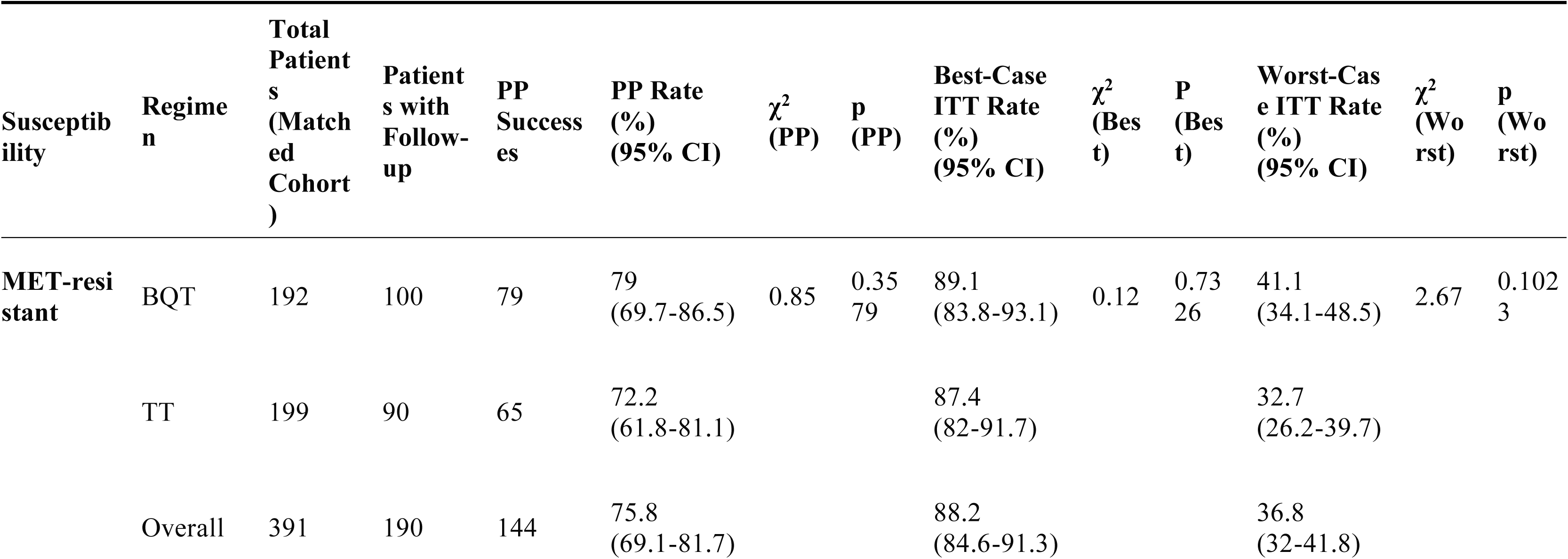
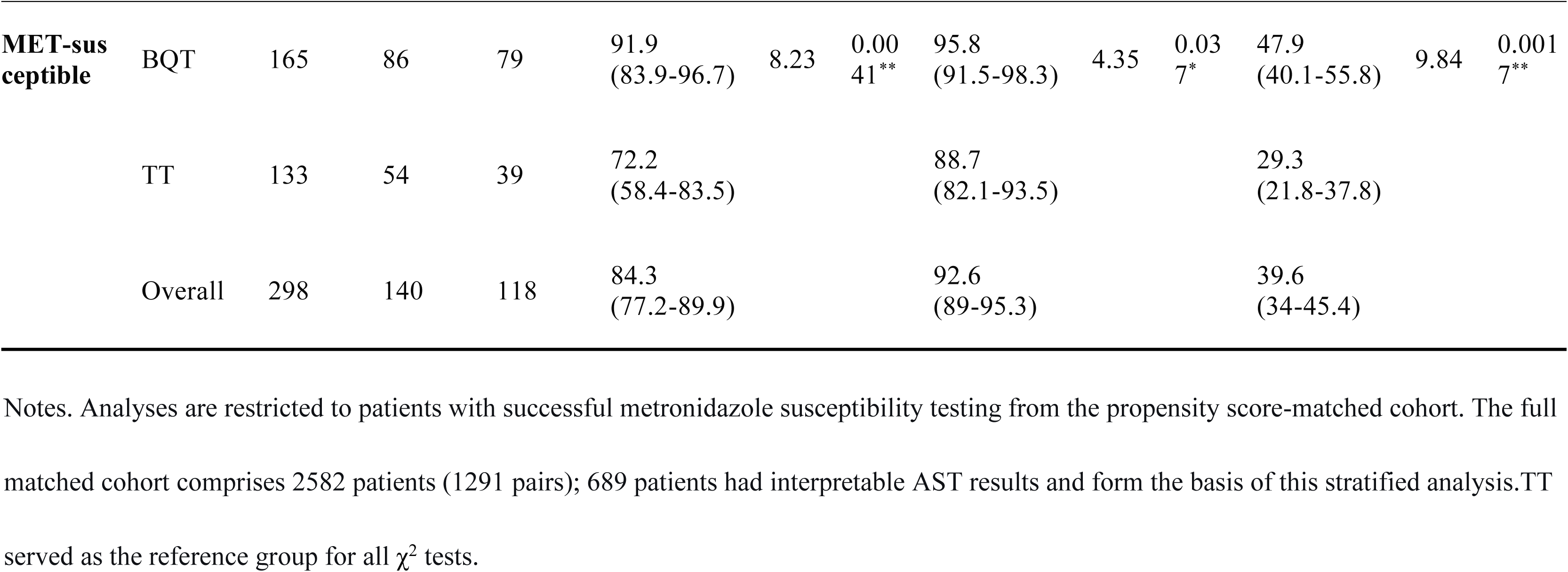
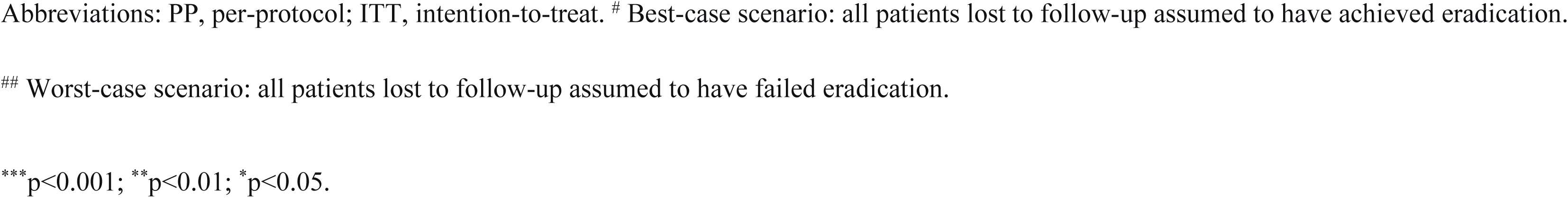
Sensitivity analysis for loss to follow-up: eradication rate estimates stratified by metronidazole susceptibility after propensity score matching (PSM).

| Susceptibility | Regimen | Total Patients (Matched Cohort) | Patients with Follow-up | PP Successes | PP Rate (%) (95% CI) | $\chi^2$ (PP) | p (PP) | Best-Case ITT Rate (%) (95% CI) | $\chi^2$ (Best) | P (Best) | Worst-Case ITT Rate (%) (95% CI) | $\chi^2$ (Worst) | p (Worst) |
| --- | --- | --- | --- | --- | --- | --- | --- | --- | --- | --- | --- | --- | --- |
| **MET-resistant** | BQT | 192 | 100 | 79 | 79 (69.7-86.5) | 0.85 | 0.3579 | 89.1 (83.8-93.1) | 0.12 | 0.7326 | 41.1 (34.1-48.5) | 2.67 | 0.1023 |
|  | TT | 199 | 90 | 65 | 72.2 (61.8-81.1) |  |  | 87.4 (82-91.7) |  |  | 32.7 (26.2-39.7) |  |  |
|  | Overall | 391 | 190 | 144 | 75.8 (69.1-81.7) |  |  | 88.2 (84.6-91.3) |  |  | 36.8 (32-41.8) |  |  |
| MET-susceptible | BQT | 165 | 86 | 79 | 91.9 (83.9-96.7) | 8.23 | 0.0041** | 95.8 (91.5-98.3) | 4.35 | 0.037* | 47.9 (40.1-55.8) | 9.84 | 0.0017** |
|  | TT | 133 | 54 | 39 | 72.2 (58.4-83.5) |  |  | 88.7 (82.1-93.5) |  |  | 29.3 (21.8-37.8) |  |  |
|  | Overall | 298 | 140 | 118 | 84.3 (77.2-89.9) |  |  | 92.6 (89-95.3) |  |  | 39.6 (34-45.4) |  |  |

Notes. Analyses are restricted to patients with successful metronidazole susceptibility testing from the propensity score-matched cohort. The full matched cohort comprises 2582 patients (1291 pairs); 689 patients had interpretable AST results and form the basis of this stratified analysis. TT served as the reference group for all $\chi^2$ tests.
Abbreviations: PP, per-protocol; ITT, intention-to-treat. # Best-case scenario: all patients lost to follow-up assumed to have achieved eradication.

**Table 2.**
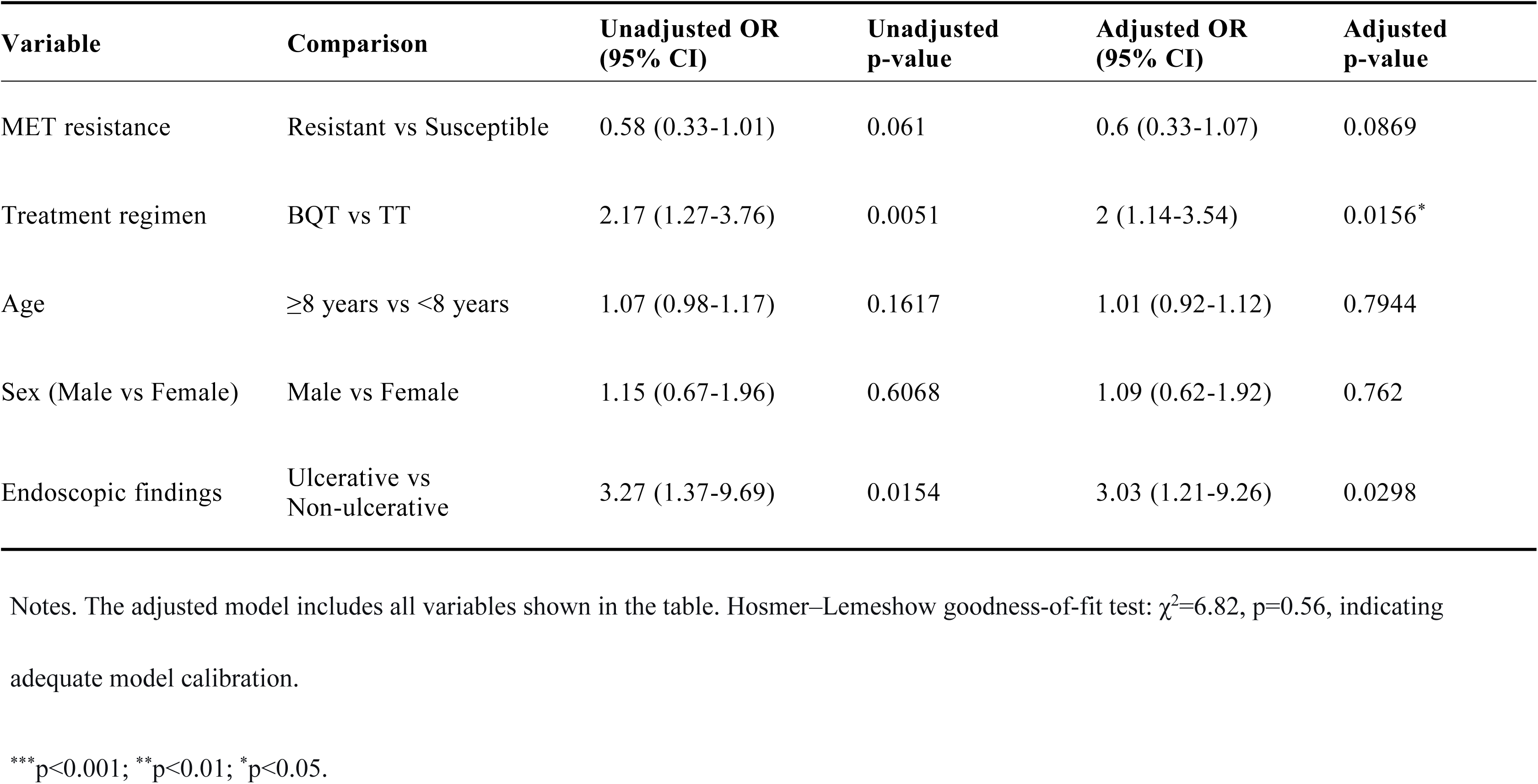
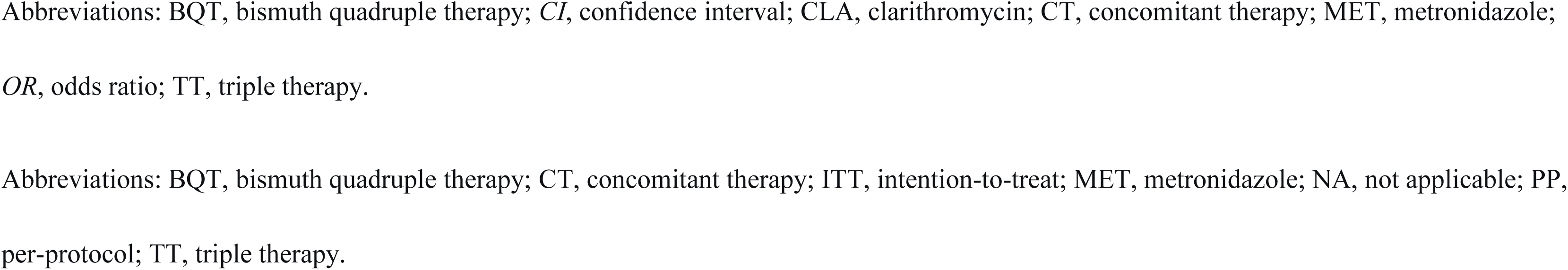
Predictors of *H. pylori* eradication success: multivariable conditional logistic regression after PSM.

| Variable | Comparison | Unadjusted OR<br>(95% CI) | Unadjusted<br>p-value | Adjusted OR<br>(95% CI) | Adjusted<br>p-value |
| --- | --- | --- | --- | --- | --- |
| MET resistance | Resistant vs Susceptible | 0.58 (0.33-1.01) | 0.061 | 0.6 (0.33-1.07) | 0.0869 |
| Treatment regimen | BQT vs TT | 2.17 (1.27-3.76) | 0.0051 | 2 (1.14-3.54) | 0.0156* |
| Age | ≥8 years vs <8 years | 1.07 (0.98-1.17) | 0.1617 | 1.01 (0.92-1.12) | 0.7944 |
| Sex (Male vs Female) | Male vs Female | 1.15 (0.67-1.96) | 0.6068 | 1.09 (0.62-1.92) | 0.762 |
| Endoscopic findings | Ulcerative vs<br>Non-ulcerative | 3.27 (1.37-9.69) | 0.0154 | 3.03 (1.21-9.26) | 0.0298 |

Notes. The adjusted model includes all variables shown in the table. Hosmer–Lemeshow goodness-of-fit test: $\chi^2=6.82$ , $p=0.56$ , indicating adequate model calibration.
\*\*\* $p<0.001$ ; \*\* $p<0.01$ ; \* $p<0.05$ .
Abbreviations: BQT, bismuth quadruple therapy; *CI*, confidence interval; CLA, clarithromycin; CT, concomitant therapy; MET, metronidazole; *OR*, odds ratio; TT, triple therapy.
Abbreviations: BQT, bismuth quadruple therapy; CT, concomitant therapy; ITT, intention-to-treat; MET, metronidazole; NA, not applicable; PP, per-protocol; TT, triple therapy.

### Checkerboard-defined additive window

All five tested strains showed additive or synergistic bismuth–MET interactions (FICI 0·375–0·75), defining the additive window at FICI≤1·0 (figure 3A). Bismuth reduced MET MIC by SF 2–32 (median 8), consistent with prior reports.^12^ MET concentrations below this window resulted in indifferent interactions, explaining the absence of bismuth benefit with low-dose MET.

**Figure 3.**
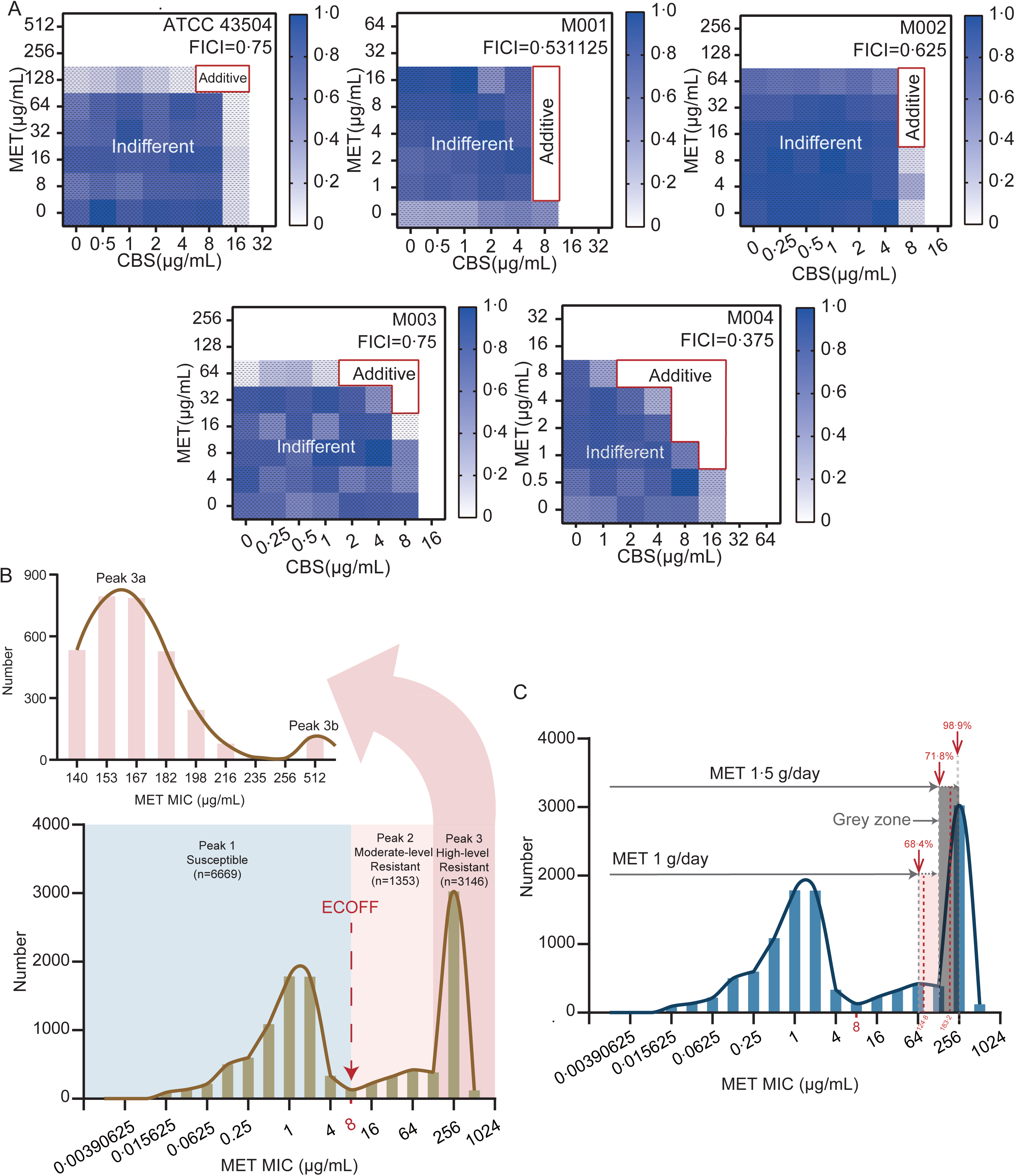
*In vitro* checkerboard assays, EUCAST MET MIC distribution, and PK/PD-predicted coverage by different MET doses. (A) Checkerboard assays of colloidal bismuth subcitrate (CBS) and MET against one reference strain (ATCC 43504) and four MET-resistant clinical isolates. The red box indicates concentration-dependent synergy (FICI≤1·0); grey area, indifference (FICI>1·0). Median synergy factor (SF)=8. FICI values are shown above each heatmap. (B) Apparent trimodal distribution of MET MIC values from the EUCAST database (n=11,168). Peak 1 (blue): susceptible population, MIC≤8 µg/mL. Peak 2 (light red): moderately resistant population, MIC range 16–128 µg/mL. Peak 3 (dark red): isolates categorised as high-level resistant by categorical susceptibility testing, with nominal MIC≥256 µg/mL. Note: Deconvolution analysis reveals that Peak 3 is itself bimodal, comprising: (1) a dominant subpopulation (Peak 3a ∼96%) with true MIC concentrated at 128–256 µg/mL within the pharmacometric grey zone; (2) a minor subpopulation (Peak 3b ∼4%) with genuine high-level resistance (true MIC>256 µg/mL). Back-calculation from clinical eradication data constrains the true geometric mean MIC of the dominant subpopulation to 150 µg/mL, substantially below the nominal 256 µg/mL breakpoint. The two-fold broth dilution method collapses the entire 128–256 µg/mL grey zone into a single ≥256 µg/mL category, thereby conflating two biologically distinct resistance subpopulations and systematically overestimating high-level resistance. Dashed red arrow indicates the EUCAST epidemiological cut-off value (ECOFF) of 8 µg/mL. (C) PK/PD-modelled coverage of the true MIC distribution with bismuth (SF=8). At 1 g/day, coverage is limited to Peaks 1+2 (68·4–71·8%); at 1·5 g/day, coverage extends into the grey zone to include ∼60% of Peak 3 (overall 71·8–98·9%). Abbreviations: CBS, colloidal bismuth subcitrate; ECOFF, epidemiological cut-off; EUCAST, European Committee on Antimicrobial Susceptibility Testing; FICI, fractional inhibitory concentration index; MET, metronidazole; MIC, minimum inhibitory concentration; PK/PD, pharmacokinetic/pharmacodynamic; SF, synergy factor.

### PK/PD coverage and grey zone quantification

At 1 g/day, MET plus bismuth (SF=8) covered isolates with MIC≤124·8 µg/mL, accounting for 68·4–71·8% of all EUCAST isolates. At 1·5 g/day, coverage extended to ∼180 µg/mL, encompassing all of Peak 2 and the majority of nominally high-resistant Peak 3 isolates, with overall population coverage of 71·8–98·9% (figure 3B, C; table 3). At 2 g/day, the coverable MIC threshold reached ≤ 244·0 µg/mL, with only a modest incremental benefit in population coverage (table 3). To reconcile ∼180 µg/mL coverage with >90% clinical eradication in MET-resistant infections,^7,8^ the dominant subpopulation of nominal Peak 3 must have true MICs below the coverage threshold. Deconvolution shows Peak 3 comprises two log-normal subpopulations: a dominant grey-zone subpopulation (Peak 3a, ∼96% of Peak 3, geometric mean MIC∼150 µg/mL) and a minor genuine high-resistance subpopulation (Peak 3b, ∼4%, geometric mean ∼512 µg/mL) (figure 3B). Under this bimodal model, 2 g/day BQT covers ∼96% of nominally high-resistant isolates (Peak 3a), and 97% of total MET-resistant isolates (Peak 2 and Peak 3).

**Table 3.**
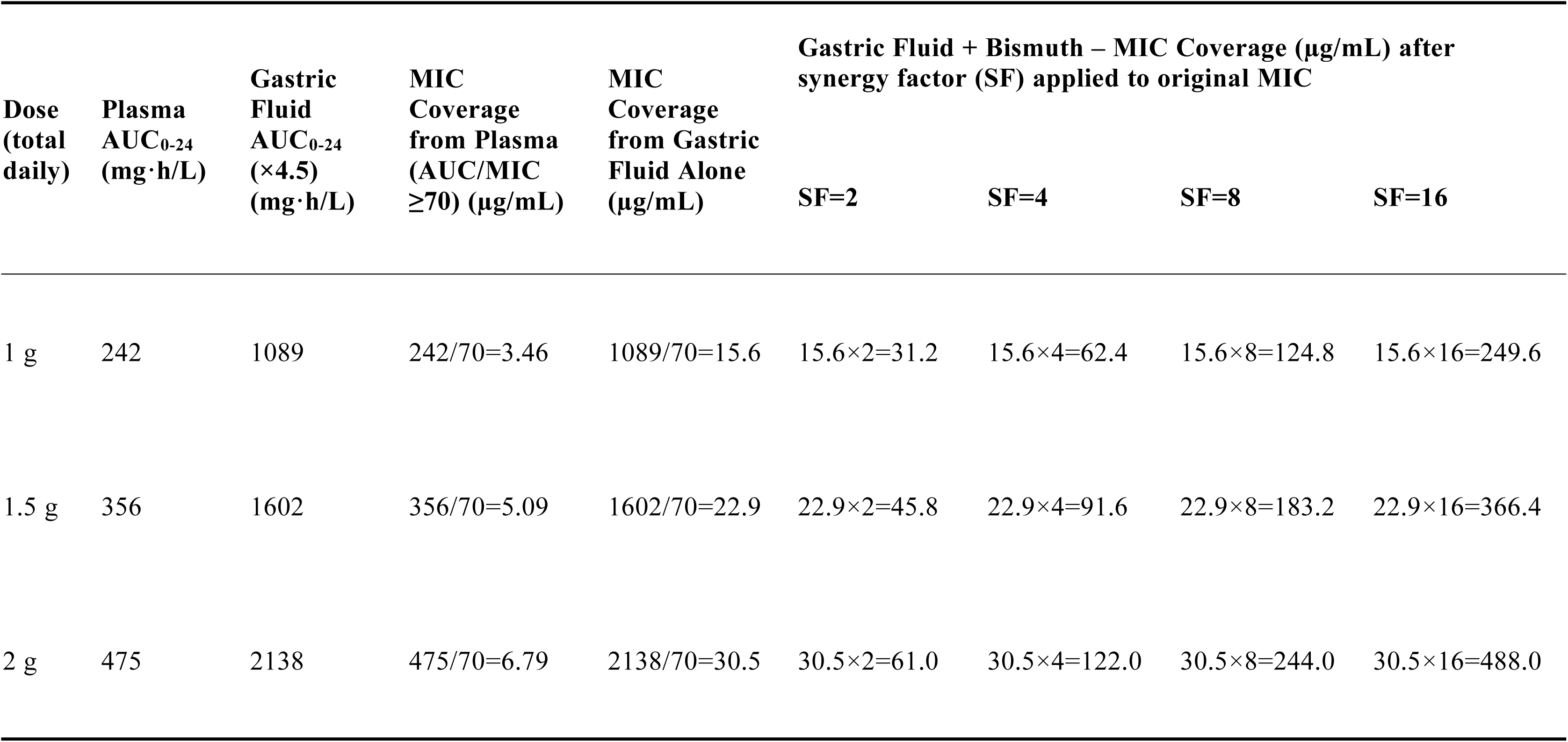
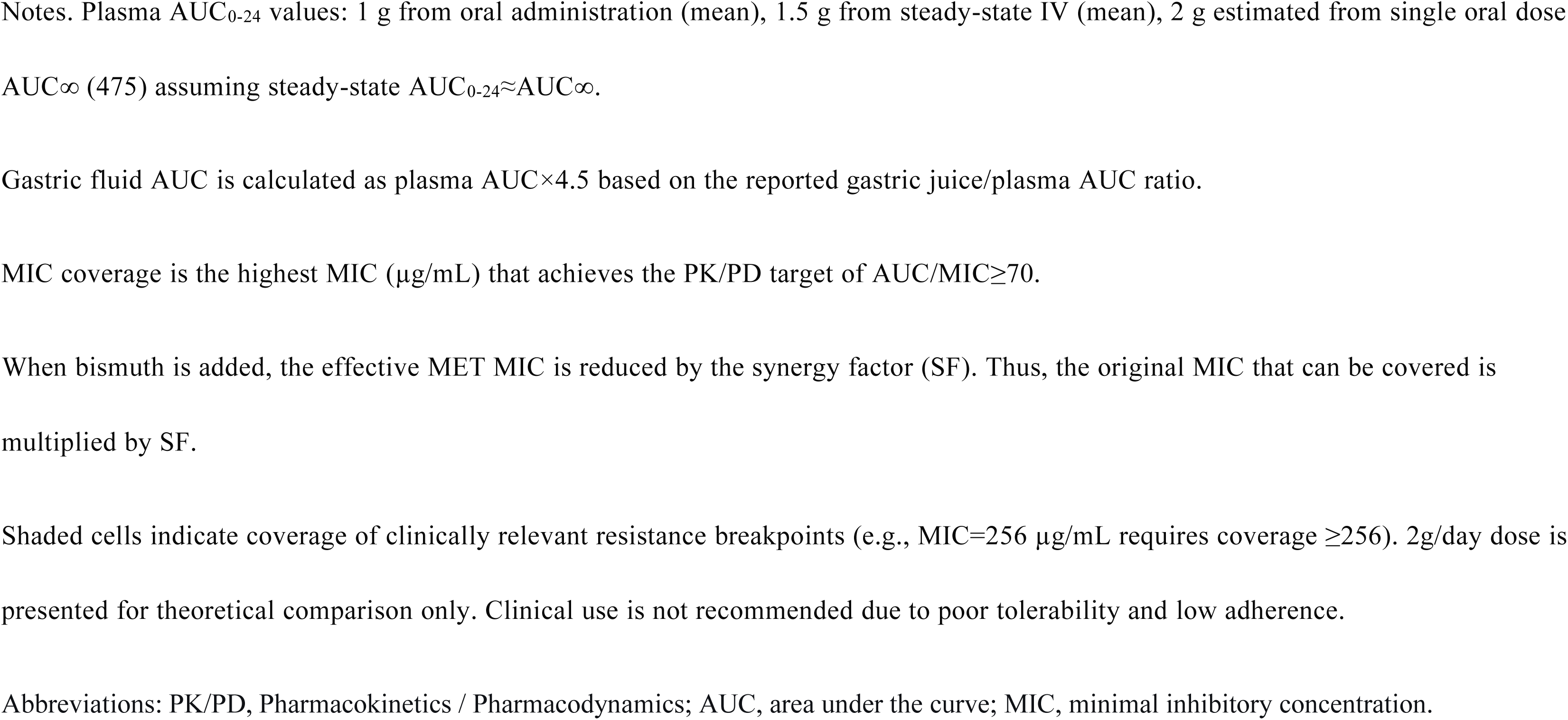
PK/PD Coverage of Metronidazole at Different Doses Alone and with Bismuth (SF=2, 4, 8, 16)

| Dose<br>(total<br>daily) | Plasma<br>AUC<sub>0-24</sub><br>(mg·h/L) | Gastric<br>Fluid<br>AUC<sub>0-24</sub><br>(×4.5)<br>(mg·h/L) | MIC<br>Coverage<br>from Plasma<br>(AUC/MIC<br>≥70) (µg/mL) | MIC<br>Coverage<br>from Gastric<br>Fluid Alone<br>(µg/mL) | Gastric Fluid + Bismuth – MIC Coverage (µg/mL) after<br>synergy factor (SF) applied to original MIC |  |  |  |
| --- | --- | --- | --- | --- | --- | --- | --- | --- |
|  |  |  |  |  | SF=2 | SF=4 | SF=8 | SF=16 |
| 1 g | 242 | 1089 | 242/70=3.46 | 1089/70=15.6 | 15.6×2=31.2 | 15.6×4=62.4 | 15.6×8=124.8 | 15.6×16=249.6 |
| 1.5 g | 356 | 1602 | 356/70=5.09 | 1602/70=22.9 | 22.9×2=45.8 | 22.9×4=91.6 | 22.9×8=183.2 | 22.9×16=366.4 |
| 2 g | 475 | 2138 | 475/70=6.79 | 2138/70=30.5 | 30.5×2=61.0 | 30.5×4=122.0 | 30.5×8=244.0 | 30.5×16=488.0 |

Notes. Plasma AUC<sub>0-24</sub> values: 1 g from oral administration (mean), 1.5 g from steady-state IV (mean), 2 g estimated from single oral dose AUC<sub>∞</sub> (475) assuming steady-state AUC<sub>0-24</sub>≈AUC<sub>∞</sub>.
Gastric fluid AUC is calculated as plasma AUC×4.5 based on the reported gastric juice/plasma AUC ratio.
MIC coverage is the highest MIC (µg/mL) that achieves the PK/PD target of AUC/MIC≥70.
When bismuth is added, the effective MET MIC is reduced by the synergy factor (SF). Thus, the original MIC that can be covered is multiplied by SF.
Shaded cells indicate coverage of clinically relevant resistance breakpoints (e.g., MIC=256 µg/mL requires coverage ≥256). 2g/day dose is presented for theoretical comparison only. Clinical use is not recommended due to poor tolerability and low adherence.
Abbreviations: PK/PD, Pharmacokinetics / Pharmacodynamics; AUC, area under the curve; MIC, minimal inhibitory concentration.

Sensitivity analyses confirmed the grey zone requires SF≥6; we used SF=8 as a conservative estimate, supported by published data showing higher SF in more resistant isolates (supplementary figure S2).^12^

## Discussion

This study provides a quantitative, hypothesis-generating framework for the clinical–microbiological discordance of MET resistance in BQT. All model-derived values are reported with full transparency of assumptions (Supplementary Methods); precision reflects computational exactitude, not empirical certainty.

We propose a concentration-dependent bismuth–MET synergy threshold: below the additive window, bismuth adds no benefit against resistant isolates; above it (achieved at ∼1·5 g/day MET), most nominally high-resistant isolates remain pharmacodynamically treatable. This aligns with >90% eradication rates reported in both adult trials and registries.^7,8^

### Three-tier strategy for MET susceptibility testing

Since 69·9% of MET-resistant EUCAST isolates fall within the obscured Peak 3,^9^ and optimised BQT covers most, routine pre-treatment MET susceptibility testing loses clinical utility. Theoretically, the resolution of MIC determination could be further refined by reducing the dilution factor or adopting fixed incremental step sizes. However, in accordance with ISO 20776-1, MIC assays carry an inherent reproducibility limit of ±1 two-fold dilution step. As such, refined measurement approaches suffer from poor reproducibility and limited practical applicability. We propose a hypothesis-generating three-tier framework:

1. Tier 1 (Treatment-naïve patients): Empiric optimised BQT (1·5 g/day MET) is recommended. Categorical MIC testing cannot distinguish treatable from untreatable resistance in this population, and high clinical efficacy renders results actionably irrelevant.
2. Tier 2 (First-line BQT failures): Categorical AST retains value to exclude alternative failure mechanisms (adherence, PPI dosing, clarithromycin resistance). A resistant MET result signals need for salvage regimens.
3. Tier 3 (Research-enriched population): Patients with MIC≥256 µg/mL who fail optimised BQT form a signal-enriched cohort. Genomic (*rdxA/frxA* profiling), metabolomic (nitroreductase activity surrogates), and SF analyses may establish predictive biomarkers that circumvent phenotypic AST entirely.^5,17^

### Safety and risk–benefit

A 14-day course of 1·5 g/day MET (21 g cumulative) is well below durations associated with clinically significant neurotoxicity.^18^ Discontinuation rates attributable to MET in modern BQT are ∼1·4%,^19–21^ and this risk is outweighed by efficacy gains. A paediatric dose of 30 mg/kg/day (equivalent to 1·5 g/day in adults) is recommended to reach the synergistic threshold.

### Limitations

1. BQT is a four-drug regimen; SF derived from clinical data may include contributions from pH effects or second-antibiotic synergy.^22,23^ However, consistency between in vitro (median SF=8) and clinical back-calculated estimates (SF≈5–11) supports bismuth–MET interaction as the dominant component.^7,8^
2. This was a single-centre retrospective study with 60% loss to follow-up. Patients lost to follow-up had a lower prevalence of ulcerative endoscopic findings (15·8% vs 19·5%) and were more likely to receive BQT (61·4% vs 54·0%) than those with complete follow-up. Given that ulcerative disease is independently associated with higher eradication success and BQT outperforms TT, differential loss to follow-up could introduce selection bias and skew efficacy estimates in either direction. While antimicrobial resistance profiles were balanced between groups and best-/worst-case ITT sensitivity analyses consistently supported the dose threshold conclusion, residual confounding cannot be excluded.
3. Multiple PK/PD model assumptions affect the precision of numerical estimates, but do not undermine the qualitative conclusion of systematic overestimation of resistance in the grey zone. These include: SF characterised in a small panel of 5 strains; the gastric-juice-to-plasma AUC ratio derived from a single study under proton pump inhibitor therapy; absence of formal Monte Carlo simulation; and use of steady-state intravenous AUC data for the 1·5 g/day reference dose. Although MET has near-complete oral bioavailability such that intravenous exposure closely approximates oral steady-state levels,^18,24^ this approximation may marginally overestimate actual intragastric concentrations and thus the achievable MIC coverage threshold.
4. Findings were derived in children, but PK/PD targets are exposure-based and age-independent, and external adult validation supports generalisability.^23^

### Future directions

Prospective validation should focus on optimised BQT failure rate as the primary endpoint, stratifying patients with MIC≥256 µg/mL by treatment outcome. Core assays would include whole-genome sequencing, targeted metabolomics, and standardised SF determination. Gastroretentive sustained-release MET formulations could further enhance intragastric exposure while reducing systemic toxicity.^25,26^

## Conclusion

We propose that bismuth–MET synergy requires a concentration-dependent additive window, and the 128–256 µg/mL interval constitutes a structural blind spot of two-fold dilution—not temporary imprecision, but a defining feature of phenotypic AST. These hypothesis-generating findings suggest routine MET susceptibility testing is of limited value for treatment-naïve patients receiving optimised BQT. Instead, we recommend restricting categorical testing to BQT failures and pursuing mechanism-based biomarkers in this enriched population to build predictive models beyond the MIC paradigm.

## Acknowledgments

We thank all children and their guardians who participated in this retrospective cohort study, as well as the clinical staff of the Department of Gastroenterology, Children’s Hospital of Fudan University for providing clinical database resources to support this research. We sincerely appreciate Professor Hong Lu, Professor Francis K. L. Chan and Professor Jyhming Liou for their valuable suggestions on research framework and manuscript revision. Special thanks to Professor Hongbin Sun for his insightful advice regarding pharmacokinetic and pharmacodynamic analytical approaches. We are also grateful to Professor Graham and Professor Dore for continuous academic encouragement throughout this research.

## Author Contributions

Chunmeng He: Conceptualization, Data curation, Formal analysis, Investigation, Methodology, Visualization, Writing – original draft, Writing – review & editing, Supervision, Project administration.

Huan Wang: Conceptualization, Methodology, Writing – review & editing.

Xinbo Xu: Methodology, Writing – review & editing.

All authors approved the final version and agree to be accountable for all aspects of the work.

## Funding

This research received no specific grant from any public, commercial, or not-for-profit funding agency.

## Conflicts of Interest

The authors declare no competing financial interests or personal relationships that could have influenced the work reported in this paper.

## Ethical Statement

The study was approved by the Institutional Ethics Committee of Children’s Hospital of Fudan University (approval No. [2017]186) and was conducted in accordance with the Declaration of Helsinki. Written informed consent was obtained from the guardians of all participants.

## Patient and Public Involvement Statement

Patients or the public were not involved in the design, conduct, reporting, or dissemination plans of this research.

## Data Availability Statement

The data supporting the findings of this study are available from the corresponding author upon reasonable request. The data are not publicly available due to privacy and ethical restrictions.

---

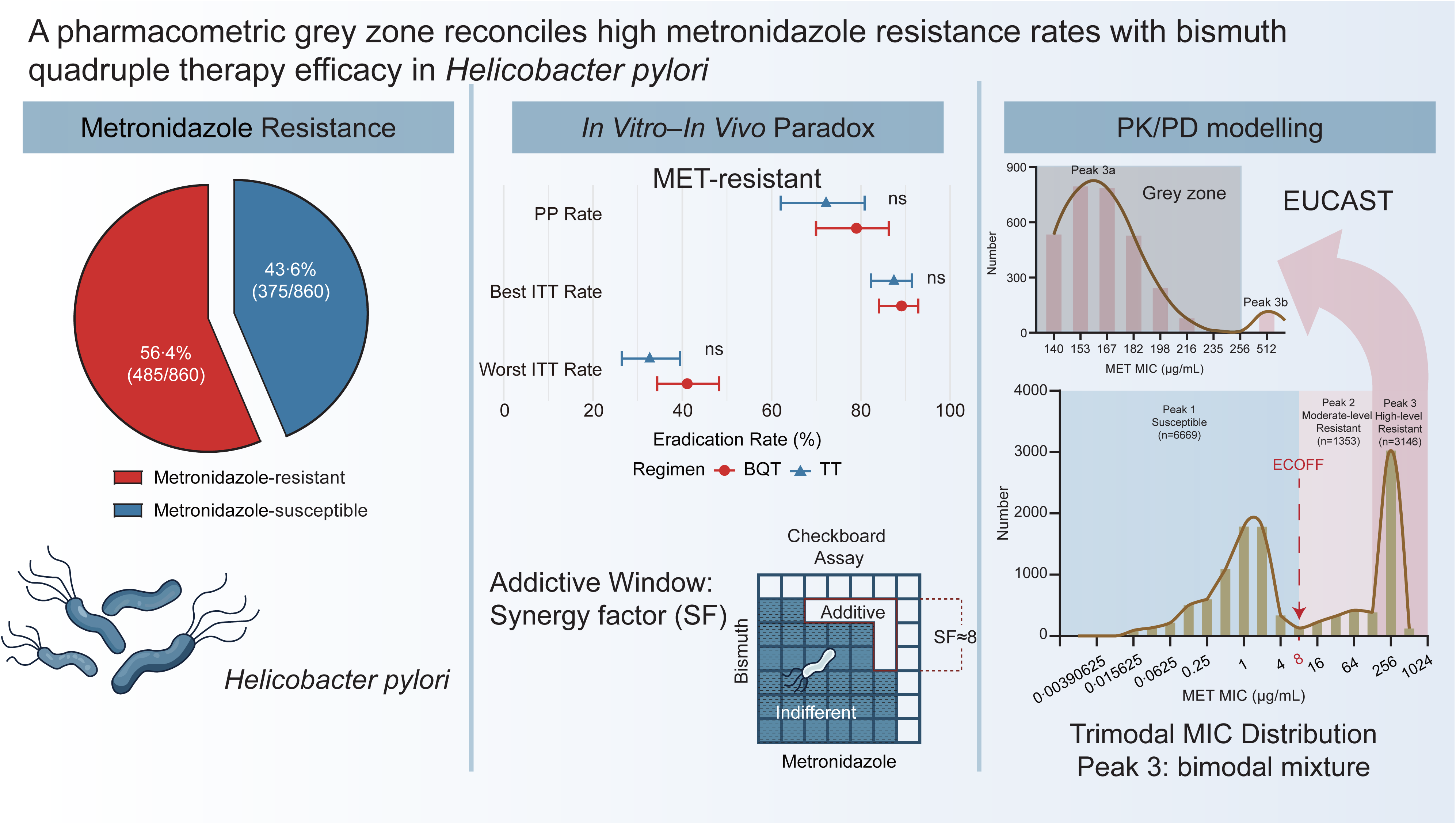

## Notes

### Competing Interest Statement

The authors have declared no competing interest.

### Author Declarations

The study was approved by the Institutional Ethics Committee of Children's Hospital of Fudan University (approval No. [2017]186) and was conducted in accordance with the Declaration of Helsinki

