## Supplementary for "A pharmacometric grey zone reconciles high metronidazole resistance rates with bismuth quadruple therapy efficacy in *Helicobacter pylori*"

#### Supplementary Methods

##### 1. Study Design and Setting

This single-centre, retrospective cohort study was conducted at the Children's Hospital of Fudan University (Shanghai, China), a National Children's Medical Centre and tertiary referral centre representing clinical practice in eastern China. Reporting followed the STROBE statement.

All patients aged 6–18 years who underwent their first oesophagogastroduodenoscopy (EGD) for digestive symptom evaluation between 1 January 2019 and 31 December 2024 were screened.

###### 1.1. Inclusion criteria

First-episode active *Helicobacter pylori* (*H. pylori*) infection diagnosed according to the 2023 ESPGHAN/NASPGHAN guidelines,[1] confirmed by at least one of:

(1) Positive culture from gastric biopsy;

(2) Concurrent positive histopathology and rapid urease test (RUT);

(3) Discordant histopathology/RUT with positive urea breath test (UBT) or stool antigen test (SAT);

(4) For bleeding peptic ulcer: single positive RUT or histopathology;

(5) First-episode eradication therapy initiated immediately after diagnosis at our centre.

##### 1.2. Exclusion criteria

(1) Incomplete clinical or microbiological data;

(2) Prior *H. pylori* eradication therapy;

(3) Use of antibiotics, PPIs, or bismuth within 4 weeks before endoscopy;

(4) Confirmed immunodeficiency;

(5) Chronic gastrointestinal comorbidities (inflammatory bowel disease, coeliac disease, congenital malformations).

#### 2. Data Collection

Data were extracted from the hospital's standardised endoscopy reporting system and electronic medical records. Pre-specified variables included:

(1) Demographics: age, sex;

(2) Clinical/endoscopic: presenting symptoms, endoscopic findings (non-ulcerative erosion vs. ulcerative erosion);

(3) Laboratory: RUT, histopathology, culture, antimicrobial susceptibility testing (AST);

(4) Treatment/outcome: regimen details, adverse events, eradication outcome.

#### 3. Ethical Approval

The study was approved by the Institutional Ethics Committee of Children's Hospital of Fudan University (approval No. [2017]186) and conducted in accordance with the

Declaration of Helsinki. Written informed consent for endoscopy and use of de-identified data for research was obtained from legal guardians at initial presentation.

#### 4. Treatment Regimens and Dosing

All patients received a standard 14-day eradication course according to the 2022 Chinese paediatric consensus,[2] without dose adjustment or early termination. Eradication was assessed ≥4 weeks after treatment completion, following a 2-week washout of PPIs, bismuth, antibiotics, and gastric mucosal protective agents.

Weight-based dosing is detailed in Supplementary Table S1. Both bismuth potassium citrate and colloidal bismuth subcitrate are collectively referred to as CBS in the manuscript.[3]

Regimens evaluated:

(1) Bismuth quadruple therapy (BQT): PPI (omeprazole)+CBS+two antibiotics

– CBS+amoxicillin (AMO)+metronidazole (MET)+OME

– CBS+clarithromycin (CLA)+AMO+OME

– CBS+CLA+MET+OME

(2) Bismuth-free concomitant therapy (CT): OME+AMO+MET+CLA

(3) Standard triple therapy (TT): OME+two antibiotics

– AMO+MET+OME

– AMO+CLA+OME

– CLA+MET+OME

#### 5. Eradication Outcome Definition

Eradication success: negative UBT or negative RUT from paired gastric biopsies ≥4 weeks after completing the 14-day course, following a 2-week washout.[1]

Eradication failure: positive result on either test at follow-up.

#### 6. *H. pylori* Culture and AST

All procedures followed EUCAST standard operating procedures.[4]

##### 6.1. Specimen Collection and Culture

Gastric biopsies (antrum and corpus) were inoculated onto Columbia blood agar within 30 min and incubated at 37 °C under microaerophilic conditions (5% $O_2$, 10% $CO_2$, 85% $N_2$) for 72–120 h.

##### 6.2. Strain Identification

Suspected colonies were identified by Gram staining (curved, seagull-shaped, or S-shaped Gram-negative bacilli) and confirmed by positive urease, catalase, and oxidase tests. Isolates were subcultured or stored at −80 °C.

##### 6.3. AST by Etest

AST was performed using the Etest method. A 0·5 McFarland suspension of pure *H. pylori* was spread onto Columbia blood agar, and Etest strips (BIO-KONT) for AMO,

CLA, MET, and levofloxacin (LEV) were applied. Plates were incubated at 37 °C under microaerophilic conditions for 72 h. MICs were read as the lowest concentration completely inhibiting visible growth.

EUCAST clinical breakpoints (v14.0, 2024):[4]

(1) AMO: >0·125 μg/mL

(2) CLA: >0·25 μg/mL

(3) MET: >8 μg/mL

(4) LEV: >1 μg/mL (surveillance only; LEV was not used therapeutically)

**7. Checkerboard Assay for CBS–MET Interaction**

The interaction between CBS (HY-B0796, MedChemExpress) and MET (HY-B0318, MedChemExpress) was tested against *H. pylori* ATCC 43504 and four clinical MET-resistant isolates.

Frozen isolates were subcultured on Columbia blood agar at 37 °C under microaerophilic conditions for 72 h. Bacterial suspensions were adjusted to ~$1\times10^6$ CFU/mL in sterile saline, then diluted 1:10 in Brucella broth+10% FBS to achieve ~$1\times10^5$ CFU/well.

Two-fold serial dilutions of CBS and MET were combined in 96-well plates in a checkerboard pattern (final volume 100 μL/well), with growth and sterility controls included. Plates were incubated at 37 °C under microaerophilic conditions for 72 h.

Growth was assessed visually and by $OD_{600}$. The MIC of each drug alone and in combination was recorded. The fractional inhibitory concentration index (FICI) was calculated as:

FICI=(MIC of CBS in combination / MIC of CBS alone)+(MIC of MET in combination / MIC of MET alone)

Interpretation:[5]

(1) Synergistic: FICI≤0·5

(2) Additive: 0·5<FICI≤1·0

(3) Indifferent: 1·0<FICI≤2·0

(4) Antagonistic: FICI>2·0

All assays were performed in duplicate on three independent occasions.

##### 7.1. Definition of the Additive Window

The additive window was defined as the range of CBS–MET concentration combinations yielding FICI≤1·0 (i.e., additive or synergistic). Below this window, the interaction becomes indifferent (FICI>1·0), and bismuth fails to potentiate MET activity against resistant strains. The lower boundary of this window informed subsequent pharmacokinetic/pharmacodynamic (PK/PD) modelling to determine the minimum intragastric MET exposure required for clinical synergy.

#### 8. Pharmacokinetic/Pharmacodynamic (PK/PD) Modelling

##### 8.1. Model Framework

PK/PD modelling was performed to simulate MET concentrations in plasma and gastric fluid and to estimate achievable MIC coverage with different MET doses, with or without bismuth synergy. MET is a concentration-dependent antimicrobial; the PK/PD index best correlated with bactericidal efficacy is the 24-hour area under the concentration–time curve to MIC ratio ($AUC_{0\text{-}24}$/MIC). A target of $AUC_{0\text{-}24}/MIC \geq 70$ was used, based on established data for anaerobic infections and consistent with previous PK/PD studies of MET against *H. pylori*.

##### 8.2. Plasma and Gastric Fluid AUC Estimation

Three total daily MET doses were simulated: 1 g, 1·5 g, and 2 g, administered once daily. Corresponding steady-state plasma $AUC_{0\text{-}24}$ values were obtained from published population pharmacokinetic studies:

(1) 1 g (oral): mean 242 mg·h/L;

(2) 1·5 g (intravenous, steady-state): mean 356 mg·h/L;

(3) 2 g (assuming linear dose-proportional pharmacokinetics and steady-state $AUC_{0\text{-}24} \approx AUC_{\infty}$): 475 mg·h/L.[6,7]

Gastric fluid $AUC_{0\text{-}24}$ was calculated as plasma $AUC_{0\text{-}24} \times 4{\cdot}5$, the reported gastric-juice-to-plasma AUC ratio,[5] reflecting marked gastric MET concentration via ion trapping.

##### 8.3. MIC Coverage

Without Bismuth For each dose, the MIC coverage achievable by MET alone was:

$$MIC_{coverage(alone)} = \frac{\text{Gastric fluid } AUC_{0-24}}{70}$$

This represents the highest MIC (μg/mL) inhibitable with ≥90% probability at the target AUC/MIC ratio.

##### 8.4. Synergy Factor (SF) and MIC Coverage With Bismuth

The synergy factor (SF) was defined as the fold-reduction in MET MIC in the presence of sufficient CBS. In our checkerboard assays, SF ranged from 2 to 32, with a median of 8, consistent with an independent external study (Andersen et al., n=42, median SF=8).[8] With bismuth, the effective MET MIC is reduced by SF; therefore, the original MIC that can be covered is multiplied by SF:

$$MIC_{coverage\ (with\ bismuth)} = \frac{\text{Gastric fluid } AUC_{0-24}}{70} \times SF$$

Sensitivity analyses used SF=2, 4, 8, and 16, spanning the additive-to-synergistic range observed *in vitro*.

##### 8.5. Comparison With Trimodal MIC Distribution

Predicted MIC coverage (alone and with bismuth) was compared with the trimodal MIC distribution of MET-resistant *H. pylori* from EUCAST surveillance data and our own AST results (n=11,168 isolates):

(1) Peak 1 (susceptible): MIC≤8 μg/mL

(2) Peak 2 (moderate resistance): MIC 16–128 μg/mL

(3) Peak 3 (high-level resistance): MIC≥256 μg/mL.[9]

##### 8.5.1. Reconciliation of Modelled Coverage with Clinical Eradication Rates

To resolve the apparent discrepancy between model-predicted MIC coverage and observed clinical eradication rates >90% in MET-resistant infections treated with 1·5 g/day BQT, we performed a quantitative analysis of the true MIC distribution within Peak 3.

###### 8.5.1.1. Required Coverage Calculation

MET-resistant isolates eradication rate=$(P_2 \times C_2)+(P_3 \times C_3)$

Where:

$P_2$, $P_3$=proportion of MET-resistant isolates in Peak 2, and 3, respectively

$C_2$, $C_3$=coverage rate for each peak For the 1·5 g/day BQT regimen (MIC coverage limit=183·2 μg/mL):

$C_2$=100% (all Peak 2 isolates have MIC ≤ 128 μg/mL < 183·2 μg/mL)

$P_2$=30·1% (proportion of isolates in Peak 2)

$P_3$=69·9% (proportion of isolates in Peak 3)

To achieve an MET-resistant isolates eradication rate >90%, we solved for the minimum required coverage within Peak 3 ($C_3$):

$90\%=(30{\cdot}1\%\times100\%)+(69{\cdot}9\%\times C_3)$

$C_3=\frac{90\%-30{\cdot}1\%}{69{\cdot}9\%}=85{\cdot}7\%$

Thus, at least 85·7% of Peak 3 isolates must have a true MIC≤183·2 μg/mL to reconcile modelled coverage with clinical observations.

##### 8.5.1.2. Log-Normal Distribution Modelling of Peak 3

MICs Consistent with standard practice in antimicrobial susceptibility research, we assumed that the natural logarithm of MIC values within Peak 3 follows a normal distribution (i.e., MIC values follow a log-normal distribution).

Let $Y=\log_2(\text{MIC})$, then $Y\sim N(\mu, \sigma^2)$, where:

$\mu=\log_2(\text{GM})$, and GM is the geometric mean MIC of Peak 3

$\sigma$=standard deviation of $\log_2$-transformed MIC values, expressed in doubling dilution steps

Based on the narrow genetic basis of high-level metronidazole resistance (complete *rdxA* inactivation with variable *frxA* impairment), the within-subpopulation standard deviation is expected to be substantially smaller than the overall cross-population value.

##### 8.5.1.3. Derivation of Constrained Geometric Mean MIC

Given the constraint that 85·7% of Peak 3 isolates have true MIC≤183·2 μg/mL (i.e., within the grey zone below the 256 μg/mL categorical breakpoint), and that all Peak 3 isolates have true MIC>128 μg/mL (the lower bound of the grey zone), we calculated the corresponding geometric mean MIC under a left-truncated $\log_2$-normal distribution:

$P[\log_2(128)<Y\leq\log_2(183{\cdot}2)]=0{\cdot}857$

$$\frac{\Phi[\frac{\log_2(183{\cdot}2)-\mu}{\sigma}]-\Phi[\frac{\log_2(128)-\mu}{\sigma}]}{1-\Phi[\frac{\log_2(128)-\mu}{\sigma}]}=0{\cdot}857$$

Where Φ(·) is the cumulative distribution function of the standard normal distribution. The standard normal quantile corresponding to a cumulative probability of 0·857 is $Z\approx1{\cdot}150$. Rearranging:

$$\mu=\log_2(128)+Z_{\text{truncated}}\cdot\sigma \quad \text{GM}=2^{\mu}$$

Sensitivity Analysis of Geometric Mean MIC Estimates Across Varying $\sigma$ Values

Constraint: $P(\text{MIC}\leq183{\cdot}2\ \mu\text{g/mL}|\text{MIC}>128\ \mu\text{g/mL})=0{\cdot}857$

| **$\sigma$ (log₂ scale)** | **GM (μg/mL)** | **Proportion of Peak 3** | **Within-peak coverage (≤183·2 μg/mL)** | **GM within grey zone?** |
|---|---|---|---|---|
| 0·15 | 161·0 | 96·1% | 89·2% | Yes |
| 0·20 | 153·1 | 96·1% | 89·2% | Yes |
| 0·25 | 143·9 | 96·1% | 89·1% | Yes |
| 0·30 | 133·4 | 96·3% | 89·0% | Yes |
| — | 128·0 | — | — | Grey zone |

| $\sigma$ ($\log_2$ scale) | GM (μg/mL) | Proportion of Peak 3 | Within-peak coverage (≤183·2 μg/mL) | GM within grey zone? |
|---|---|---|---|---|
| | (threshold) | | | lower bound ↓ |
| 0·35 | 121·9 | 96·4% | 88·9% | No |
| 0·40 | 109·8 | 96·5% | 88·8% | No |

This yields a constrained true geometric mean MIC of approximately 128–160 μg/mL for the high-resistance Peak 3, substantially below the nominal categorical breakpoint of 256 μg/mL and squarely within the 128–256 μg/mL pharmacometric grey zone.

##### 8.5.2. Discretized Log-Normal Modelling of True MIC Distribution in Peak 3

###### 8.5.2.1. Model Assumptions

Consistent with standard antimicrobial susceptibility modeling and the log-normal framework defined above, the natural log-transformed true MIC values within Peak 3 follow a normal distribution:

$$Y = \log_2(\text{MIC}) \sim N(\mu, \sigma^2)$$

Where:

(1) $\mu = \log_2$ (MIC): Log geometric mean of true continuous MIC for Peak 3; median constrained estimate GM=150 μg/mL ($\sigma$=0·2), derived from clinical cure constraint $P$(MIC≤183·2)=85·7% within Peak 3.

(2) $\sigma$=0·2: Standard deviation of log-transformed MIC, median value from the accepted range 0·15–0·30 for high-resistance *H. pylori* subpopulations.

(3) Discretization step: Fine geometric increment of $2^{1/8}$≈1·0905-fold (root eighth of two), generating sequential discrete MIC nodes starting at 140 μg/mL up to 256 μg/mL, consistent with continuous true MIC partitioning to resolve the hidden distribution between the two-fold dilution cutoffs (128 and 256 μg/mL) used in routine EUCAST testing.

(4) Hard population constraints from experimental susceptibility data:

1) Total isolates assigned EUCAST categorical MIC=256: $N_{140\text{–}256}$=3024

2) Total isolates assigned EUCAST categorical MIC=512 (>256): $N_{>256}$=122

3) Total Peak 3 population: $N_{\text{Peak3}}$=3024+122=3146

###### 8.5.2.2. Discrete MIC Node Generation

We constructed evenly spaced log-scale MIC nodes using a $2^{1/8}$≈1·0905 multiplicative step from the lower bound of the hidden distribution (128×$2^{1/8}$≈140 μg/mL) up to the categorical breakpoint 256 μg/mL: $MIC_{k+1}=MIC_k\times 2^{1/8}$ Generated discrete MIC grid points: 140, 153, 167, 182, 198, 216, 235, 256 μg/mL

Each value represents the central bin midpoint for log-normal probability integration over its corresponding log-MIC interval.

###### 8.5.2.3. Probability Density & Isolate Count Derivation

**Step 1: Standard normal *Z*-score transformation for each discrete MIC node**

For each discrete MIC value $m_k$:

$$Z_k = \frac{\log_2(m_k) - \mu}{\sigma}$$

Where $\mu = \log_2(150) \approx 7{\cdot}23$ , $\sigma = 0{\cdot}2$.

**Step 2: Cumulative normal probability for each MIC bin**

The proportion of isolates falling within each discrete MIC interval $[m_k, m_{k+1})$ is calculated via the standard normal cumulative distribution function $\Phi(\cdot)$:

$$P(m_k \leq \text{MIC} < m_{k+1}) = \Phi(Z_{k+1}) - \Phi(Z_k)$$

The cumulative probability of all bins from 140 to 256 yields $P(140 \leq \text{MIC} \leq 256)$; the residual upper tail probability is $P(\text{MIC} > 256) = 1 - \Phi(Z_{256})$.

**Step 3: Normalization to match observed categorical isolate counts**

Raw uncalibrated bin probabilities from the log-normal density were rescaled to strictly satisfy the experimental count constraints:

(1) Sum of raw probabilities for all bins 140–256:

$$P_{\text{raw, 140–256}} = \sum P(m_k \leq \text{MIC} < m_{k+1})$$

(2) Per-bin scaling factor for the 140–256 range:

$$SF_{\text{bin}}=\frac{3024}{N_{\text{Peak 3}}\times P_{\text{raw, 140–256}}}$$

(3) Calibrated isolate count for each discrete MIC bin k:

$$n_{\text{k}}=N_{\text{Peak 3}}\times P(m_{\text{k}}\leq\text{MIC}<m_{\text{k+1}})$$

(4) Isolates with true MIC>256 were fixed to the observed count $n_{>256}$=122, consistent with EUCAST categorization MIC=512.

**Step 4: Coverage summation for the 1·5 g/day BQT regimen**

All discrete MIC bins with midpoint ≤183·2 µg/mL (140, 153, 167, 182) were summed to calculate the total number of Peak 3 isolates covered at the regimen's MIC coverage limit:

$$N_{\text{covered}}=n_{140}+n_{153}+n_{167}+n_{182}$$

Proportion of covered isolates within the EUCAST MIC=256 categorical group:

$$C_3=\frac{N_{\text{covered}}}{3024}$$

**8.5.2.4. Final Calibrated Discrete Isolate Counts (GM=150, $\sigma$=0·2, $2^{1/8}$-fold step)**

| Discrete MIC bin (µg/mL) | Calibrated number of isolates | Covered by 1·5 g/day BQT (MIC≤183·2 µg/mL) |
|---|---|---|
| 140 | 540 | Yes |

| | | |
|---|---|---|
| 153 | 801 | Yes |
| 167 | 793 | Yes |
| 182 | 534 | Yes |
| 198 | 249 | No |
| 216 | 85 | No |
| 235 | 19 | No |
| 256 | 3 | No |
| > 256 (EUCAST MIC=512) | 122 | No |
| Total Peak 3 isolates | 3146 | — |

###### 8.5.2.5. Key Output for Clinical Reconciliation

We find that within the MIC range >128 µg/mL, the MIC distribution resolves into two distinct peaks, designated Peak 3a and Peak 3b. When combined with the MIC≤128 µg/mL data, we conclude that Peak 3 is not a continuation of Peak 2. This is supported by extrapolation of the log-normal distribution fitted to Peak 2 (16–128 µg/mL), which predicts only ~290 isolates in the 128–256 µg/mL interval, whereas

3,024 are observed—a >10-fold discrepancy that cannot be explained by tail extension of a unimodal distribution.

Total isolates within the EUCAST MIC=256 category with true MIC≤183·2 μg/mL:

$N_{covered}$=540+801+793+534=2668

Proportion of covered isolates among categorical MIC=256 strains:

$$C_{3a}=\frac{2668}{3024}=88\cdot 2\%$$

$$C_{3}=\frac{2668}{3146}=84\cdot 8\%$$

This discretized log-normal partitioning demonstrates that over 80% of isolates categorized as high-level resistant (MIC=256 μg/mL) by two-fold dilution testing have true continuous MIC values below the 183·2 μg/mL coverage threshold of the 1·5 g/day BQT regimen, resolving the apparent mismatch between categorical susceptibility results and clinical eradication rates >90%.

**8.5.2.6. Sensitivity Analysis Extension**

Identical discretization workflows can be repeated across the full log-standard deviation range ($\sigma$=0·15- 0·40) and alternative geometric step sizes ($\sqrt{2}=2^{1/2}$, $\sqrt[4]{2}=2^{1/4}$) to evaluate robustness of the covered isolate proportion to choice of fine discretization scale and log-MIC variance. All analyses retain the hard count constraints $N_{140\text{–}256}$=3024 and $N_{>256}$=122.

**8.5.3. Extended Coverage Analysis for 2 g/day Regimen**

We extended our analysis to the 2 g/day metronidazole BQT regimen, which achieves a higher MIC coverage limit of 244·0 μg/mL (gastric AUC=2,138 mg·h/L divided by the PK/PD target AUC/MIC≥70, further augmented by a bismuth SF of 8).

###### 8.5.3.1. Coverage Calculation Under the Bimodal Mixture Model

Using the same bimodal mixture model parameters derived for Peak 3 (Section 8.5.2), we calculated the proportion of isolates covered by the 2 g/day regimen:

For the Peak 3a (grey zone, $GM_{3a}$=153·1 μg/mL, $\sigma_{3a}$=0·2 $\log_2$):

The conditional probability of true MIC≤244 μg/mL, given MIC>128 μg/mL, is:

$$P(\mathrm{MIC}\leq 244 \mid \mathrm{MIC}>128)=\frac{\Phi[\frac{\log_2(244)-\mu_{3a}}{\sigma_{3a}}]-\Phi[\frac{\log_2(128)-\mu_{3a}}{\sigma_{3a}}]}{1-\Phi[\frac{\log_2(128)-\mu_{3a}}{\sigma_{3a}}]}=99\cdot 96\%$$

For the Peak 3b (genuine high-level resistance, $GM_{3b}$=512 μg/mL, $\sigma_{3b}$=0·3 $\log_2$):

$$P(\mathrm{MIC}\leq 244 \mid \mathrm{MIC}>128)=0\cdot 02\%$$

Overall coverage in Peak 3 (weighted by mixture proportions $f_{3a}$=96·1%, $f_{3b}$=3·9%):

$$C_{2g}=f_{3a}\cdot P_{3a}+f_{3b}\cdot P_{3b}=96\cdot 1\%$$

This corresponds to 3,023 out of 3,146 isolates in the nominal high-resistance category being covered by the 2 g/day regimen.

###### 8.5.3.2. Comparison with 1·5 g/day Regimen

| Regimen | Coverage threshold (μg/mL) | Overall coverage in Peak 3 | Isolates covered inPeak 3 | Overall coverage in MET-resistant isolates | Isolates covered in MET-resistant isolates |
|---|---|---|---|---|---|
| 1·5 g/day MET-containing BQT | 183·2 | 85·70% | 2,697/3,146 | 90·0% | 4,050/4,499 |
| 2·0 g/day MET-containing BQT | 244 | 96·10% | 3,023/3,146 | 97·3% | 4,376/4,499 |

Increasing the MET dose from 1·5 g/day to 2·0 g/day extends coverage by 10·4 percentage points, rescuing an additional 326 isolates from the nominal high-resistance category. The remaining ~4% of uncovered isolates belong almost exclusively to the Peak 3b with true MIC values >256 μg/mL, representing genuine high-level resistance that cannot be overcome by dose escalation alone.

###### 8.5.3.3. Sensitivity Analysis

The coverage estimate for the 2 g/day regimen is highly robust across the biologically plausible range of $\sigma_1$:

| $\sigma_1$ ($\log_2$ scale) | $GM_1$ (μg/mL) | Primary subpopulation proportion | 2 g/day coverage |
|---|---|---|---|

| | | | |
|---|---|---|---|
| 0·15 | 161 | 96·1% | 96·1% |
| 0·2 | 153·1 | 96·1% | 96·1% |
| 0·25 | 143·9 | 96·2% | 96·0% |
| 0·3 | 133·4 | 96·3% | 96·0% |

Across all tested $\sigma$ values, the 2 g/day coverage remains consistently at ~96%, confirming that the finding is not sensitive to the specific distributional assumptions for the primary subpopulation.

##### 8.6. External Clinical Validation

Two independent real-world datasets were used to back-calculate the SF required to achieve the reported eradication rates in MET-resistant infections. The SF needed to cover MIC=128 µg/mL (upper limit of Peak 2) and MIC=256 µg/mL (lower limit of Peak 3) was calculated as:

$$SF=\frac{70\times \mathrm{MIC}}{\text{Gastric fluid AUC}_{0-24}}$$

(1) Dataset 1 – Chinese adult trial:[10] BQT with MET 1·6 g/day, 94·7% eradication.

Plasma $AUC_{0\text{-}24}$≈387·2 mg·h/L; gastric AUC≈1,742·4 mg·h/L.

For MIC 128 µg/mL: SF≈5·1

For MIC 256 µg/mL: SF≈10·3.

(2) Dataset 2 – Hp-EuReg registry:[11] BQT with MET 1·5 g/day, 91% eradication.

Plasma $AUC_{0\text{-}24}$=356 mg·h/L; gastric AUC=1,602 mg·h/L.

For MIC 128 µg/mL: SF≈5·6

For MIC 256 µg/mL: SF≈11·2

The clinically effective SF required for >90% eradication lies between these values, with a median estimate of ≈8—consistent with our checkerboard-derived median SF.

##### 8.7. Assumptions and Limitations

(1) Fixed gastric/plasma AUC ratio of 4·5 (may vary with gastric pH and food intake);

(2) Linear pharmacokinetics (dose-proportional AUC);

(3) AUC/MIC target of 70 derived from non-gastric infections; a precise *H. pylori*-specific gastric target has not been validated;

(4) SF assumed independent of MET concentration within the tested range;

(5) All simulations performed using Microsoft Excel.

#### 9. Statistical Analysis

No formal sample size calculation was performed; all eligible patients over the 6-year period were included. Analyses were conducted using SPSS 20.0 (IBM Corp.). A two-sided p<0·05 was considered significant. Missing baseline data (<5%) were not imputed; complete-case analysis was used.

##### 9.1. Descriptive and Comparative Statistics

Normality was assessed by the Shapiro–Wilk test and visual inspection. Normally distributed continuous variables are presented as mean±SD and compared by Student's t-test; non-normally distributed variables as median (IQR) and compared by the Mann–Whitney U test. Categorical variables are presented as n (%) and compared by Pearson's $\chi^2$ or Fisher's exact test. Exact 95% confidence intervals (CIs) were calculated.

##### 9.2. Sensitivity Analysis for Loss to Follow-Up

The primary efficacy analysis was the per-protocol (PP) analysis in the propensity score-matched sample. To assess the robustness of the PP findings to loss to follow-up, we performed two extreme-scenario intention-to-treat (ITT) analyses:

(1) Best-case ITT: all patients lost to follow-up assumed eradicated;

(2) Worst-case ITT: all patients lost to follow-up assumed to have failed.

These analyses were conducted in the same matched sample and are reported as sensitivity analyses.

##### 9.3. Propensity Score Matching (PSM) to Address Confounding by Indication

Because treatment assignment (BQT vs. TT) was non-randomised and baseline differences existed between groups, we performed 1:1 propensity score matching on the full cohort (including both followed-up and lost-to-follow-up patients, 2,582 patients after matching; 1291 matched pairs). The propensity score was estimated using logistic regression with the following baseline covariates: age, sex, endoscopic findings (ulcerative vs. non-ulcerative erosion), and treatment year (Supplementary Figure S1). Nearest-neighbour matching was performed without replacement, using a caliper of 0·2×SD of the logit propensity score. Balance was assessed by standardised mean difference <0·1. After matching, all comparative efficacy analyses (per-protocol, best-case ITT, worst-case ITT) were conducted within the matched sample. Post-matching comparisons used McNemar's test.

A multivariable conditional logistic regression model was then built using backward stepwise elimination (removal criterion p>0·05). Adjusted odds ratios with 95% confidence intervals are reported. Model calibration was assessed using the Hosmer–Lemeshow test adapted for conditional logistic regression.

##### 9.4. Reporting of Unmatched Analyses

For transparency, unmatched intention-to-treat and per-protocol analyses in the full cohort are provided in Supplementary Tables. Because of significant confounding by indication, these unmatched results were not interpretable as causal treatment effects and are not discussed in the main manuscript.

#### Supplementary Figures

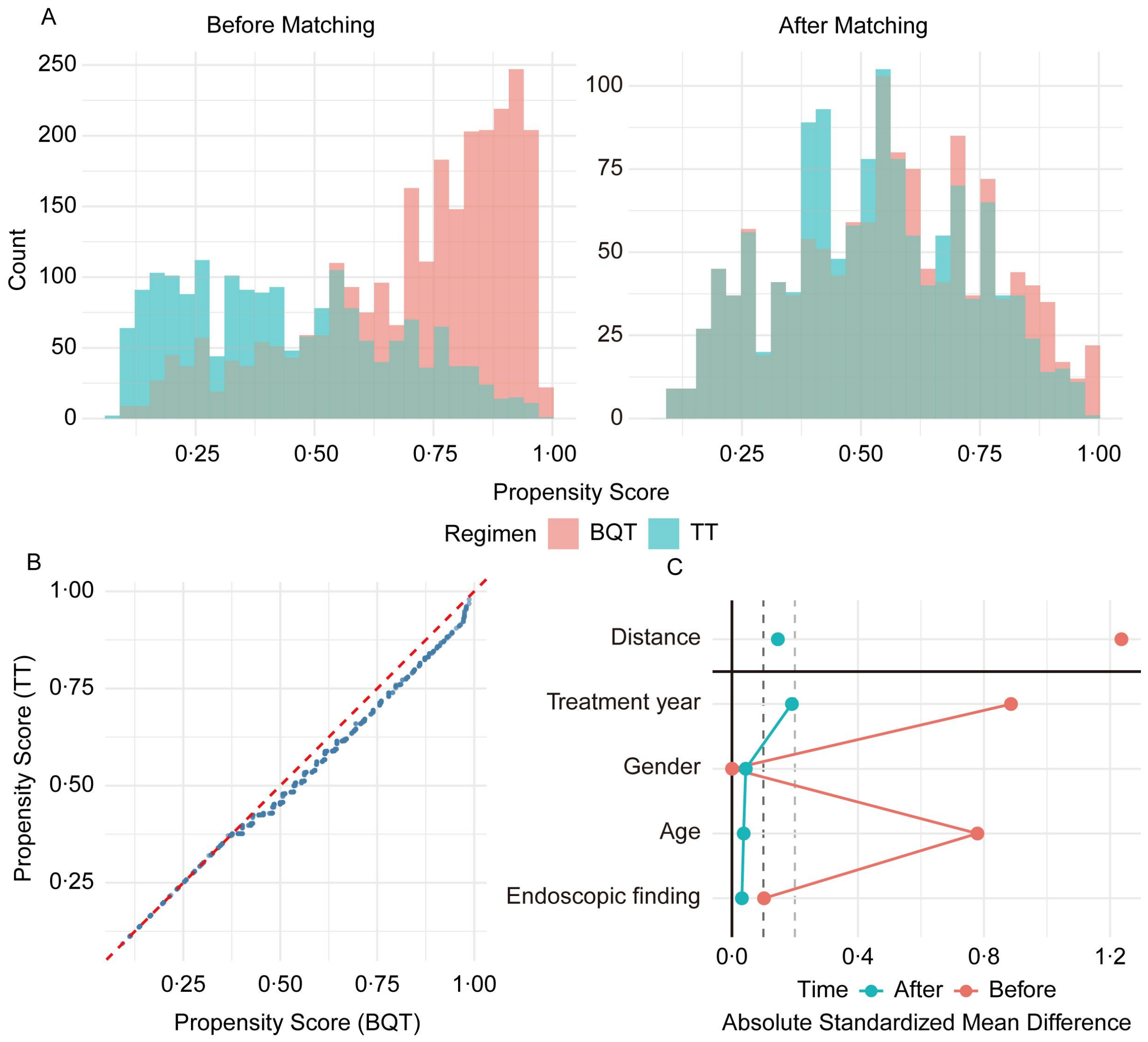

**Supplementary Figure S1. Results of propensity score matching (PSM) between the bismuth quadruple therapy (BQT) and triple therapy (TT) regimens.**

(A) Histograms displaying the distribution of propensity scores for the two treatment groups before (left panel) and after (right panel) PSM. The overlapping areas indicate improved balance in baseline covariates between the BQT and TT groups following matching.

(B) Scatter plot comparing the propensity scores of the control group against those of the treatment group, demonstrating the alignment of scores after the matching process. Points clustering along the diagonal line suggest successful matching.

(C) Love plot illustrating the absolute mean differences (standardized mean differences) of covariates between the two groups before (light red dots) and after (light blue dots) adjustment. Covariates include distance, treatment year, gender, age, endoscopic finding. The shift of blue dots closer to zero indicates a significant reduction in bias and better balance achieved after PSM.

Abbreviations: PSM, propensity score matching; BQT, bismuth quadruple therapy; TT, triple therapy; MET, metronidazole; CLA, clarithromycin.

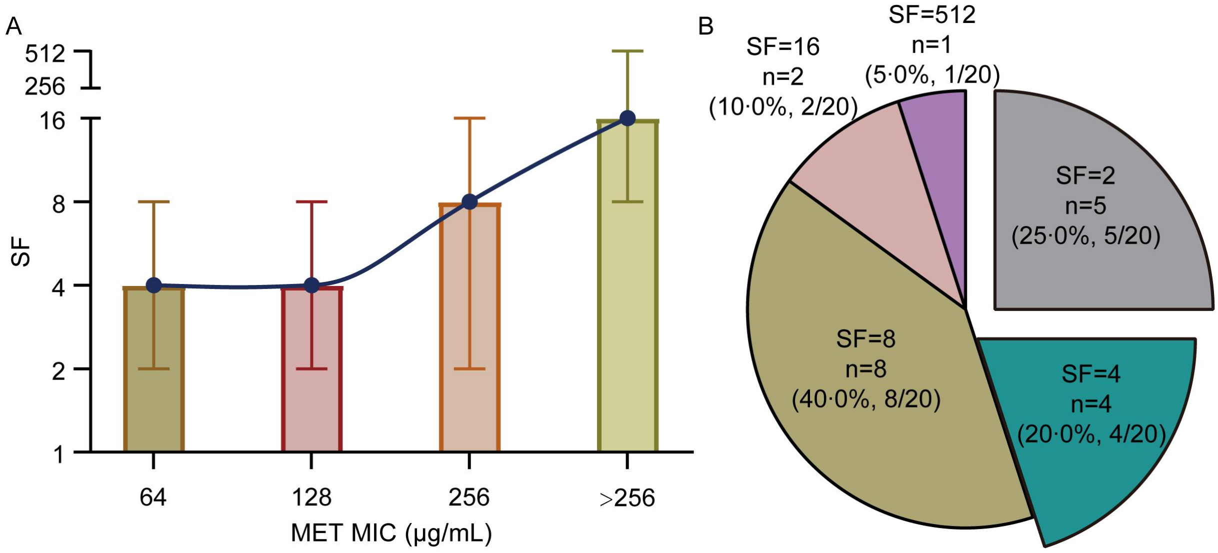

**Supplementary Figure S2. Analysis of the Synergistic Factor (SF) between bismuth and metronidazole against *H. pylori* strains with varying resistance levels.**

(A) Relationship between Metronidazole (MET) Minimum Inhibitory Concentration (MIC) and the Synergistic Factor (SF). For data processing, MIC values exceeding 256 µg/mL were standardized to 512 µg/mL. One extreme outlier (SF=5446·8) was excluded from the analysis. The trend line suggests that as MET resistance increases (higher MIC), the SF tends to increase; however, this correlation did not reach statistical significance due to the limited sample size. Data are presented as mean ± standard deviation.

(B) Distribution of SF values among MET-resistant strains (defined as MET MIC>8 µg/mL). The pie chart illustrates the proportion of strains falling into different SF categories. While 55% of all MET-resistant strains exhibited SF≥6—below the 75% threshold typically cited for robust synergistic consistency—this overall distribution is heavily influenced by lower-MIC resistant strains with weaker synergy. Among high-level resistant isolates (MIC≥256 μg/mL), SF was consistently >8 (Andersen et al., 7/8 strains; Figure S2A). Thus, SF=8 represents a conservative estimate for the Peak 3 subpopulation relevant to the grey zone hypothesis.

Abbreviations: SF, Synergistic Factor; MET, Metronidazole; MIC, Minimum Inhibitory Concentration; SD, Standard Deviation.

#### Supplementary Tables

**Supplementary Table S1. Standard weight-based dosing for paediatric *H. pylori* eradication.**

| Drug | Daily Dose | Frequency | Maximum per Dose |
|---|---|---|---|
| Omeprazole (OME) | 1·0 mg/kg | twice daily | 20 mg |
| Colloidal bismuth subcitrate (CBS)* | 6–8 mg/kg (bismuth element) | twice daily, before meals | 165 mg |
| Amoxicillin (AMO) | 50 mg/kg | twice daily | 1,000 mg |
| Metronidazole (MET) | 20 mg/kg | twice daily | 500 mg |
| Clarithromycin (CLA) | 15–20 mg/kg | twice daily | 500 mg |

* Bismuth potassium citrate (commercial name: Livzon Dele, Livzon Group)—the most widely used bismuth formulation in paediatric practice in mainland China—shares an identical molecular formula and chemical structure with colloidal bismuth subcitrate, as verified by the NCBI PubChem database, and is classified as an official synonym of CBS. The dose is expressed as elemental bismuth.

Abbreviations: AMO, amoxicillin; CBS, colloidal bismuth subcitrate; CLA, clarithromycin; MET, metronidazole; OME, omeprazole.

**Supplementary Table S2. Baseline characteristics of patients with and without post-treatment assessment**

| Characteristic | Follow-up (n=1,844) | Loss to follow-up (n=2,766) | $\chi^2$ | p |
|---|---|---|---|---|
| Gender | | | 1·14 | 0·286 |
| Male | 1,064 (57·7%; 55·4–60·0%) | 1,552 (56·1%; 54·2–58·0%) | | |
| Female | 780 (42·3%; 40·0–44·6%) | 1,214 (43·9%; 42·0–45·8%) | | |
| Age (years), mean ± SD | 9·3 ± 3·2 | 9·9 ± 3·2 | – | 0·060[†] |
| Endoscopic findings | | | 11·38 | 0·001[**] |
| Non-ulcerative erosion | 1,484 (80·5%; 78·6–82·3%) | 2,329 (84·2%; 82·8–85·5%) | | |
| Ulcerative erosion | 360 (19·5%; 17·7–21·4%) | 437 (15·8%; 14·5–17·2%) | | |
| Culture positivity[‡] | 1,320/1,756 (75·2%; 73·1–77·2%) | 1,370/2,629 (52·1%; 50·2–54·0%) | 223·63 | <0·001[***] |

| Characteristic | Follow-up (n=1,844) | Loss to follow-up (n=2,766) | $\chi^2$ | p |
|---|---|---|---|---|
| AST success | 551/1,320 (41·7%; 39·1–44·4%) | 309/1,370 (22·6%; 20·4–24·9%) | 113·46 | <0·001*** |
| Any resistance§ | 351/551 (63·7%; 59·6–67·7%) | 196/309 (63·4%; 57·9–68·6%) | 0·01 | 0·936 |
| AMO resistance | 16/551 (2·9%; 1·8–4·7%) | 7/309 (2·3%; 1·1–4·6%) | 0·32 | 0·572 |
| CLA resistance | 144/551 (26·1%; 22·6–30·0%) | 86/309 (27·8%; 23·0–33·1%) | 0·29 | 0·589 |
| LEV resistance | 2/551 (0·36%; 0·10–1·3%) | 2/309 (0·65%; 0·18–2·3%) | 0·36 | 0·551 |
| MET resistance | 309/551 (56·1%; 51·9–60·2%) | 176/309 (57·0%; 51·4–62·4%) | 0·06 | 0·806 |
| Treatment regimen | | | | |
| BQT | 995 (54·0%; 51·7–56·3%) | 1,699 (61·4%; 59·6–63·2%) | 25·39 | <0·001*** |
| CBS+AMO+MET+PPI | 556/995 (55·9%; 52·7–59·0%) | 1,030/1,699 (60·6%; 58·3–63·0%) | 5·26 | 0·022* |

| Characteristic | Follow-up (n=1,844) | Loss to follow-up (n=2,766) | $\chi^2$ | p |
|---|---|---|---|---|
| CBS+CLA+AMO+PPI | 144/995 (14·5%; 12·3–16·8%) | 268/1,699 (15·8%; 14·1–17·6%) | 0·82 | 0·365 |
| CBS+CLA+MET+PPI | 295/995 (29·6%; 26·8–32·6%) | 401/1,699 (23·6%; 21·6–25·7%) | 11·97 | 0·001** |
| CT | 44 (2·4%; 1·7–3·2%) | 65 (2·4%; 1·8–3·0%) | 0·02 | 0·888 |
| TT | 805 (43·6%; 41·4–46·0%) | 1,002 (36·2%; 34·4–38·0%) | 25·62 | <0·001*** |
| AMO+MET+PPI | 60/805 (7·5%; 5·7–9·5%) | 65/1,002 (6·5%; 5·0–8·2%) | 0·65 | 0·421 |
| AMO+CLA+PPI | 612/805 (76·0%; 72·9–78·9%) | 780/1,002 (77·8%; 75·1–80·4%) | 0·84 | 0·361 |
| CLA+MET+PPI | 133/805 (16·5%; 14·0–19·3%) | 157/1,002 (15·7%; 13·5–18·1%) | 0·24 | 0·623 |

Notes. Data are presented as n (%; 95% CI) unless otherwise indicated. Continuous variables are expressed as mean ± SD and compared using Student's t-test. Pearson's $\chi^2$ test was used; Yates' continuity correction was applied when expected frequencies were <5, and Fisher's exact test was used when expected frequencies were <1.

† p value from Student's t-test for independent samples.

‡ Culture positivity rates are based on patients who underwent gastric mucosal culture (follow-up: n=1,756; loss to follow-up: n=2,629). Denominators differ from the overall cohort because culture was not performed or could not be completed in all patients.

§ Resistance rates are based on isolates with successful antimicrobial susceptibility testing (follow-up: n=551; loss to follow-up: n=309). A global $\chi^2$ test was used to compare overall resistance profiles between groups.

[***]p<0.001; [**]p<0.01; [*]p<0.05.

Abbreviations: AMO, amoxicillin; CLA, clarithromycin; MET, metronidazole; LEV, levofloxacin; BQT, bismuth quadruple therapy; CT, concomitant therapy; TT, triple therapy; CBS, colloidal bismuth subcitrate; PPI, proton pump inhibitor.

**Supplementary Table S3. Sensitivity analysis for loss to follow-up: eradication rate estimates stratified by metronidazole susceptibility full cohort, unmatched.**

| **Susceptibility** | **Regimen** | **Total Patients (Full Cohort)** | **Patients with Follow-up** | **PP Successes** | **PP Rate (%) (95% CI)** | **$\chi^2$ (PP)** | **p (PP)** | **Best-Case ITT Rate (%) (95% CI)** | **$\chi^2$ (Best)** | **p (Best)** | **Worst-Case ITT Rate (%) (95% CI)** | **$\chi^2$ (Worst)** | **p (Worst)** |
|---|---|---|---|---|---|---|---|---|---|---|---|---|---|
| **MET-resistant** | BQT | 468 | 188 | 158 | 84·0 (78·0–88·9) | 6·22 | 0·014* | 93·6 (91·1–95·5) | 8·60 | 0·003** | 33·8 (29·7–38·1) | 0·00 | 0·947 |
| | TT | 247 | 116 | 84 | 72·4 (63·6–79·9) | | | 87·0 (82·4–90·7) | | | 34·0 (28·3–40·1) | | |
| | Overall | 715 | 304 | 242 | 79·6 (74·7–83·8) | | | 91·3 (89·1–93·3) | | | 33·8 (30·5–37·3) | | |
| **MET-susceptible** | BQT | 330 | 113 | 102 | 90·3 (83·5–94·5) | 14·41 | <0·001*** | 96·7 (94·3–98·2) | 21·23 | <0·001*** | 30·9 (26·1–36·2) | 0·05 | 0·831 |
| | TT | 198 | 91 | 63 | 69·2 (59·0–77·9) | | | 85·9 (80·2–90·2) | | | 31·8 (25·8–38·5) | | |

| Susceptibility | Regimen | Total Patients (Full Cohort) | Patients with Follow-up | PP Successes | PP Rate (%) (95% CI) | $\chi^2$ (PP) | p (PP) | Best-Case ITT Rate (%) (95% CI) | $\chi^2$ (Best) | p (Best) | Worst-Case ITT Rate (%) (95% CI) | $\chi^2$ (Worst) | p (Worst) |
|---|---|---|---|---|---|---|---|---|---|---|---|---|---|
| | Overall | 528 | 204 | 165 | 80·9 (74·9–85·8) | | | 92·6 (90·1–94·6) | | | 31·3 (27·5–35·3) | | |

Notes. TT served as the reference group for all $\chi^2$ tests.

### Best-case scenario: all patients lost to follow-up assumed to have achieved eradication.

#### Worst-case scenario: all patients lost to follow-up assumed to have failed eradication.

***p<0·001; **p<0·01; *p<0·05.

Abbreviations: BQT, bismuth quadruple therapy; CT, concomitant therapy; ITT, intention-to-treat; MET, metronidazole; NA, not applicable; PP, per-protocol; TT, triple therapy.
